# Simulation-Trained Deep Learning for Automated Cell-Based HLA Antibody Assay Interpretation in Pre-Transplant Diagnostics

**DOI:** 10.64898/2026.07.23.26358785

**Authors:** Cassian Afting, Anna-Lena Semmler, Salim Oulghazi, Julia I. Ries, Anouk E. Lichtenberg, Julius FE Jaeger, Christina Merk, Daniel Fürst, Hanns-Martin Lorenz, Torsten Tonn, Christian Seidl, Tarik Exner

## Abstract

Preformed and *de novo* antibodies against donor human leukocyte antigen (HLA) antigens remain a major cause of antibody-mediated rejection and graft loss after organ transplantation. Although solid-phase assays and virtual crossmatching have reshaped pre-transplant risk assessment, physical crossmatching, which assesses whether recipient antibodies react with donor cells, remains widely used as the final compatibility assessment before transplantation. These workflows include both complement-dependent cytotoxicity (CDC) and flow cytometry crossmatch (FCXM) assays, but their interpretation remains partly manual, operator-dependent, and, for microscopic CDC readout, semi-quantitative. Here, we present AlloViewer, a web-based software platform for automated and traceable interpretation of image-based and flow-cytometry-based HLA antibody diagnostics. For CDC microscopy, AlloViewer employs a simulation-trained deep learning (UNet) workflow that combines automated lymphocyte segmentation with experiment-specific fluorescence classification and well-level cytotoxicity scoring. To deliberately capture the technical variability encountered in routine diagnostics, we generated simulated CDC-like training images spanning differences in image resolution, acquisition conditions, staining quality, cell density, cell distribution, clustering, and background fluorescence, among others. The resulting simulation-trained model enabled robust lymphocyte detection across heterogeneous imaging conditions and outperformed a conventional rule-based image analysis pipeline under variable acquisition conditions. Automated CDC scoring achieved performance within the range of human inter-annotator variability and approached the practical reproducibility limit defined by human disagreement. To cover all modalities of pre-transplant physical crossmatching, AlloViewer further supports automated FCXM interpretation through cell population identification and population-specific immunoglobulin G (IgG) readouts. The platform integrates these workflows in an assay-specific web interface and provides application programming interface (API) access for programmatic submission of assay data and retrieval of processed results. Together, AlloViewer establishes an integrated computational framework for standardized and traceable interpretation of CDC crossmatch assays, CDC-based HLA antibody identification testing using commercial test-cell panels, and FCXM workflows. More broadly, this work demonstrates how simulation-trained artificial intelligence (AI) can facilitate robust computational analysis across technically heterogeneous laboratory environments, providing a framework for standardizing traditionally operator-dependent diagnostic workflows.

## Introduction

Preformed and *de novo* antibodies against donor human leukocyte antigen (HLA) antigens remain a major cause of antibody-mediated rejection and graft loss after transplantation^1,2^. Accordingly, current guidelines emphasise systematic HLA antibody screening, definition of unacceptable antigens, and final assessment of donor-recipient compatibility by virtual and/or physical crossmatch, particularly in kidney transplantation ^3–9^. Solid-phase assays, such as Luminex single-antigen bead testing, and virtual crossmatching have transformed pre-transplant risk assessment and reduced the centrality of cell-based assays in many programmes^3,4,7,8,10^. However, physical crossmatching by cell-based complement-dependent cytotoxicity (CDC) or flow cytometry crossmatch (FCXM) remains widely used, either as a mandatory core or supplemental pre-transplant recipient-donor compatibility method, and thus continues to provide an important assay-level assessment of antibody reactivity against donor cells. While solid-phase antibody testing and virtual crossmatching increasingly produce standardized and digitally traceable outputs, interpretation of physical crossmatch results often remains more dependent on manual read-out and expert judgement. This creates a need for objective, reproducible, and auditable interpretation workflows.

This need is particularly apparent for CDC-based screening and crossmatching, which are still performed in a substantial number of European Federation for Immunogenetics (EFI)-, American Society for Histocompatibility and Immunogenetics (ASHI)-, and British Society for Histocompatibility and Immunogenetics (BSHI)-accredited histocompatibility laboratories worldwide, and remain embedded in external proficiency schemes as well as in specific clinical or resource-limited settings^3,4,8,11–13^. Microscopic CDC read-out, however, is still manual and semi-quantitative. Technologists usually visually estimate the proportion of lysed versus viable lymphocytes in wells that may contain hundreds to thousands of cells, and then map these estimates onto ordinal scores^14^. This process is regularly time-consuming and inherently subjective^15^, with particular variability around decision thresholds, such as approximately 20% lysed cells, where small differences in perceived positivity can change the categorical result of the entire diagnostic. Contamination of lymphocyte preparations with other nucleated blood cells further complicates interpretation, as these cells often need to be mentally excluded while still contributing fluorescence signal. Although guidelines discuss analytical pitfalls, including immunoglobulin M (IgM) and non-HLA reactivity, and recommend enhanced methods, they offer limited practical means to standardise microscopic read-out beyond score tables and expert oversight. As a result, CDC remains one of the least objectively quantified components of the modern HLA testing repertoire, even where it is routinely performed.

In contrast, deep learning-based image analysis, and UNet-like convolutional architectures in particular, continue to become the de facto standard for cell and tissue segmentation across microscopy modalities^16–19^. These approaches reliably delineate individual cells in dense fields, enable accurate cell counting and classification, and have already entered diagnostic workflows in digital pathology and other areas^20,21^. Yet, they have not been applied to the routine microscopic CDC diagnostics: there are no tools that automatically segment and classify individual lymphocytes in CDC assays and return per-well positivity fractions and quality metrics suitable for direct use in a diagnostic histocompatibility laboratory.

In FCXM, comparable but more post-analytical challenges arise during the interpretation of raw cytometry data^22,23^. Although FCXM provides a more quantitative read-out than microscopic CDC scoring, data analysis is still frequently based on visual expert assessment, including manual population identification and interpretation of population-specific fluorescence shifts. This has been identified as a relevant source of interlaboratory variability^24–26^. Computational methods for objective, reproducible cytometry analysis have therefore been proposed as an important means of standardisation^27^. Yet despite increasing adoption of algorithm-based gating and interpretation in other diagnostic and research settings, their use has not reached pre-transplant crossmatching.

In this study, we present AlloViewer, an automated software platform designed to standardize the interpretation of cell-based HLA antibody diagnostics by integrating computational image analysis and flow cytometry interpretation within a traceable diagnostic workflow. For CDC-based assays, AlloViewer combines simulation-trained deep learning for robust lymphocyte detection with experiment-specific fluorescence classification to generate objective well-level cytotoxicity measurements and quality metrics directly from fluorescence microscopy images. To ensure robustness across heterogeneous laboratory imaging conditions, model development was based on simulated image data spanning a broad range of acquisition settings, staining characteristics, cell densities, clustering patterns, and background fluorescence, among others. The platform further extends to FCXM analysis by supporting automated cell population detection and population-specific immunoglobulin G (IgG) signal quantification from raw flow cytometry data. Together, these components establish an integrated computational framework for standardized, traceable, and reproducible interpretation of CDC crossmatching, CDC-based HLA antibody identification testing, and FCXM workflows in pre-transplant diagnostics.

## Materials and Methods

### Isolation of Lymphocytes from Peripheral Blood and Spleen

For lymphocyte isolation from peripheral whole blood, 2 mL of blood was mixed with 80 µL of RosetteSep™ Human Total Lymphocyte Enrichment Cocktail (Stemcell Technologies, Cat.: #15263) and incubated for 20 min at room temperature (RT). For experiments requiring isolation of specific lymphocyte subsets, 80 µL of RosetteSep™ Human T Cell Enrichment Cocktail (Stemcell Technologies, Cat.: #15061) or RosetteSep™ Human B Cell Enrichment Cocktail (Stemcell Technologies, Cat.: #15064) was used instead of the total lymphocyte cocktail, while all subsequent steps were performed identically. Subsequently, 2 mL of washing solution (Dulbecco’s Phosphate Buffered Saline (PBS; Sigma-Aldrich, Cat.: #D8537) supplemented with 2% Fetal Bovine Serum (FBS; PAN-Biotech, Cat.: #P30-3301) and 0.372 g/L EDTA (PanReac AppliChem, Cat.: #A3553,0250)) was added, and the mixture was slowly layered onto the density gradient solution Lymphodex (InnoTrain, Cat.: #002041500). The tube was centrifuged at 1369 g for 15 min with the brake turned off. The separated lymphocytes were collected, washed with washing solution, and centrifuged twice, first at 270 g for 10 min and then at 152 g for an additional 10 min. The washed cells were resuspended in McCoy’s 5A Medium (Park and Terasaki modification; Gibco, Cat.: #80014020) containing 0.5% FBS, with or without 0.01 M 1,4-dithiothreitol (DTT; Carl Roth, Cat.: #6908.1). DTT conditions were used to control for IgM antibody-mediated positivity of the respective reactions.

For lymphocyte isolation from spleen tissue, the tissue was incised multiple times with a scalpel, and cells were flushed out using a syringe in RPMI-1640 (Roswell Park Memorial Institute Medium; InnoTrain, Cat.: #002043500). The resulting cell suspension was passed through a sieve layered with a gauze pad. A 3 mL aliquot (or 2 mL and 1 mL whole blood) of the filtrate was then mixed with 120 µL of the respective RosetteSep™ Cocktail (Human Total Lymphocyte, Human T Cell, or Human B Cell Enrichment Cocktail; Stemcell Technologies, Cat.: #15263, #15061, and #15064, respectively), and the isolation procedure was performed identically to the protocol used for lymphocyte isolation from peripheral whole blood from this step onward.

For flow cytometric crossmatch experiments, lymphocytes were isolated separately from anticoagulated whole blood by magnetic negative selection as described below.

### Complement-Dependent Cytotoxicity (CDC) Assay

For CDC crossmatches, unloaded microtitre plates (Greiner Bio-One, Cat.: #653190) were first loaded with paraffin oil droplets (Carl Roth, Cat.: #8904.2). Isolated lymphocyte suspensions in McCoy’s 5A Medium (Park and Terasaki modification) supplemented with 0.5% FBS were transferred at a volume of 1 µL into each well, and 1 µL of patient or control serum was added. The plate was incubated for 30 min at RT. Next, 5 µL of reconstituted, lyophilized rabbit complement mix (BAG Diagnostics, Cat.: #7018) was added to each well and incubated for a further 60 min at RT. Subsequently, 5 µL of FluoroQuench (OneLambda, Thermo Fisher Scientific, Cat.: #FQAC500) was added to each well, incubated for 30 min at 4°C in the dark, and finally subjected to epifluorescence microscopy.

For DTT conditions, patient sera and IgM control sera were preincubated in 5 mM DTT for 30 min at 37°C, after which 1 µL of L-cystine (4.8 mg/mL in PBS; pH 7.4; Sigma-Aldrich, Cat.: #C8755) was added and incubated for 5 min prior to the addition of the complement solution.

For the identification of HLA class I antibodies and characterization of their specificities, the SeraScreen Abs microtitre plate kit (BAG Diagnostics, Cat.: #7252) was used according to the manufacturer’s instructions. Briefly, cell-containing plates were thawed at 37°C, loaded with prewarmed 20 µL of Terasaki Park Medium (BAG Diagnostics, Cat.: #7028) supplemented with 10% FBS, and incubated for 15 min at RT. The medium was then removed, and 2 µL of negative control, positive control, and patient serum were added to the respective wells, followed by incubation for 30 min at RT. The remaining steps of the assay were performed identically to the protocol used for the CDC crossmatch from this step onward.

### Flow Cytometric Crossmatch (FCXM)

For flow cytometric crossmatches (FCXM) we followed a modified version of the Halifaster FCXM protocol described in detail elsewhere^28^. In that, lymphocytes were isolated from EDTA- or citrate phosphate dextrose adenine (CPDA)-anticoagulated whole blood by negative magnetic selection using the EasySep™ Direct Human Total Lymphocyte Isolation Kit (Stemcell Technologies, Cat.: #19655). Briefly, 1.5-5 mL whole blood was transferred into 15 mL round-bottom tubes and incubated per 1 mL blood with 50 µL isolation cocktail and 50 µL magnetic particles for 5 min at room temperature (RT). Samples were then diluted with an equal volume of Dulbecco’s phosphate-buffered saline without calcium and magnesium (DPBS; Gibco, Cat.: #14190-094), placed on an EasyEights™ EasySep™ Magnet (Stemcell Technologies, Cat.: #18103) for 5 min, and the lymphocyte-containing supernatant was transferred to a fresh tube. A second magnetic depletion step with the same amount of magnetic particles was performed, followed by two additional 5 min magnetic separations to further reduce non-lymphocytic contaminants. The isolated cells were pelleted, resuspended in DPBS, and counted in a Neubauer chamber (C-Chip Neubauer improved; Carl Roth, Cat.: #PK36.1) after 1:10 dilution in Türk’s solution (Sigma-Aldrich, Cat.: #1.09277.0100). All isolation steps were performed at RT.

Before FCXM analysis, patient sera were centrifuged for 10 min at 10000 g to remove aggregates. Isolated lymphocytes were treated with Pronase (Merck, Cat.: #53702-10KU) for 15 min at 37 °C, washed twice with FACS wash buffer (FWB; DPBS supplemented with 2% fetal calf serum (FCS; Biological Industries, Cat.: #04-400-1A)), and adjusted to 1 × 10^7 cells/mL in DPBS. For each reaction, 15 µL cell suspension (1.5 × 10^5 cells) and 30 µL serum were incubated in U-bottom 96-well plates for 20 min at RT on a shaker. Each run included a secondary-antibody-only control using DPBS, a negative serum pool, a strong positive serum pool, a weak positive control generated by approximately 1:32 dilution of the positive pool with the negative pool, duplicate patient-serum wells, and a 1:5 dilution of patient serum in DPBS to assess a prozone effect. Cells were then washed three times with FWB by centrifugation for 2 min at 500 g and stained for 10 min at RT in the dark with a master mix containing CD3-PerCP (clone SK7; BD Biosciences, Cat.: #345766), CD19-APC-Cy7 (clone HIB19; BioLegend, Cat.: #302218), and a fluorescein isothiocyanate (FITC)-conjugated F(ab’)_2_ goat anti-human immunoglobulin G (IgG) Fcγ fragment-specific secondary antibody (Jackson ImmunoResearch, Cat.: #109-096-098). After two rounds of final washes, cells were resuspended in 250 µL FWB and acquired on a FACSLyric flow cytometer (BD, Becton Dickinson GmbH, Heidelberg, Germany) equipped with 405, 488, and 640 nm lasers and operated at medium flow rate (60 µL/min). The instrument underwent daily Performance Quality Control (QC) using CS&T beads before acquisition. At least 10,000 events were recorded per well.

### Fluorescence Microscopy and Imaging

Fluorescence microscopy was performed using FluoroQuench staining, containing Ethidium Bromide (excitation peak: 301 nm, emission peak: 603 nm) and Acridine Orange (excitation peak: 490 nm, emission peak: 520 nm). Color epifluorescence imaging was performed on an Olympus CKX53 microscope using a 4×/0.10 objective for microscope-camera acquisition and a 10×/0.25 objective for smartphone-based acquisition. On this system, both fluorophores were illuminated simultaneously at their respective excitation wavelengths, and the resulting emission signal was collected using a long-pass filter. Color images were acquired using an LC35 Color USB3 camera (Evident Scientific) mounted via a U-TV0.5XC-3-8 0.5× adapter. Images were acquired at a resolution of 2160 × 1620 px with the following camera settings: exposure 200 ms, white balance R: 1.47, G: 1, B: 1.5, sharpness 5, gamma 1, saturation 1, and brightness 0. Images were saved and further used in .tiff format.

Additional monochrome fluorescence imaging was performed on a Keyence BZ-X810 microscope using a 10×/0.30 PlanFluor objective. In contrast to the simultaneous color epifluorescence imaging on the Olympus system, monochrome images were acquired using separate fluorescence filter sets for DAPI, GFP, and TRITC detection, with excitation/emission bandpass filters of 360/40 nm and 460/50 nm, 470/40 nm and 525/40 nm, and 545/25 nm and 605/70 nm, respectively. Images were acquired with an exposure time of 1/30 s at a resolution of 1920 × 1440 px as 8-bit images.

For smartphone-based imaging, an iPhone XR and Google Pixel 9 were attached to one ocular of the Olympus CKX53 microscope using a NexYZ Smartphone Adapter (Celestron). Images were acquired with default camera settings and subsequently used as input images in the native .jpg format for the GooglePixel device or converted from .heic to .jpg format for the iPhone XR.

### Synthetic image generation and camera-domain adaptation

Synthetic CDC assay images were generated in two separate steps. First, a stochastic scene simulator created RGB images with a circular well, fluorescent cells, optical artifacts, and exact pixel-level targets. Second, a camera-style module transformed the rendered scene to resemble specific acquisition domains while leaving all labels unchanged.

Synthetic fluorescence microscopy images were generated within an explicit circular well geometry with random center jitter, radial background intensity, vignette, wall glow, optional ring artifacts, and fine/coarse background texture. Cell labels, diameters, brightness, blur, ellipticity, and rotation were sampled per image or per cell. Cell diameters were drawn from image-size-calibrated bounds, with a legacy scaling model retained as fallback.

Cells were positioned inside the well with optional rim enrichment, one-sided bias, and explicit cluster placement. Clusters could form either elongated chain-like structures or compact packed groups, followed by KD-tree-based overlap reduction while keeping cells within the well. Final instance masks were generated from the settled cell geometry, and cells were rendered from these masks so that image signal and ground-truth labels shared the same shape. Positive and negative cells were rendered with jittered orange-like and green-like fluorescence signals, respectively. “Ghost” cells outside the main focus-plane of the well, debris, wall reflections, and ring artifacts were added as image-only structures and excluded from ground-truth labels.

The simulator returned the RGB image, metadata, and exact supervision targets consisting of all inside-well cell instances and the corresponding binary cell mask; auxiliary outputs could additionally include boundary and focus-filtered audit targets.

After scene rendering, camera-domain adaptation was applied to the RGB image only. Device presets were defined for a three-channel microscope, monochrome microscope, iPhone XR, Google Pixel, and unmodified synthetic images (for exact details refer to the camera descriptions above). The camera model sampled domain-specific parameters for exposure, per-channel gain and offset, contrast, brightness, white balance, saturation, channel mixing, uneven illumination, vignette, tone mapping, gamma, blur, sharpening, read noise, resizing artifacts, clipping, and JPEG compression. To align synthetic images with real acquisition statistics, reference images from each device domain were used to estimate per-channel RGB quantile distributions. Foreground-like and background-like regions in real images were estimated semi-automatically using thresholding by the Otsu method (for full implementation details refer to the source code). Synthetic images were then adjusted by monotone quantile remapping, using the simulated cell mask to apply separate foreground and background mappings with a soft transition between them. A final optional channel-wise median correction shifted synthetic channel medians toward the target device domain.

Camera realism was checked by visual inspection, RGB histograms, and PCA of image-level appearance features from real and synthetic images. Synthetic datasets were stored as HDF5 files containing RGB images, instance labels, target maps, and JSON metadata with sampled simulation and camera parameters. Depending on the dataset mode, scenes were padded and resized, cropped around the well, tiled, or stored at full resolution. Each training tile contained four supervision channels: cell mask, soft boundary map, center heatmap, and energy map. Full implementation details are given in the provided source code.

### UNet architecture and configurations

A family of UNet models was used for 2-D segmentation with the same architecture and three channel capacities for web deployment. Inputs were RGB tensors with shape N × 3 × H × W, where N is the batch size. Outputs were raw logits with shape N × Cout × H × W from a final 1 × 1 convolution, where Cout is configurable, for example for multiple target maps such as interior and boundary maps, or for a single mask.

Each convolutional block used two depthwise-separable convolution sequences. Each sequence consisted of a 3 × 3 depthwise convolution followed by a 1 × 1 pointwise convolution, GroupNorm, and ReLU activation. Optional spatial dropout was applied after the first sequence in each block. The encoder consisted of an initial convolutional block followed by four 2 × 2 max-pooling operations, reducing the spatial resolution to approximately H/16 × W/16 at the bottleneck. To reduce computation, the bottleneck kept the channel width at 8 × base_ch rather than expanding further.

The decoder used bilinear upsampling by a factor of two, skip connections by channel concatenation, and padding of the upsampled feature maps where needed to match the corresponding encoder feature maps for odd spatial sizes. The final prediction therefore matched the input spatial size. Three model sizes differed only in the base channel width: small used base_ch = 16, medium used base_ch = 32, and large used base_ch = 64. With Cout = 4, these corresponded to approximately 106k, 398k, and 1.54M trainable parameters, respectively. Channels doubled along the encoder up to 8 × base_ch and were reduced again along the decoder. Convolutional weights were initialized with He normal initialization. Normalization scale parameters were initialized to one and biases to zero.

### UNet training and loss functions

We trained UNet models with three RGB input channels and four output channels predicting the cell mask, boundary map, center probability map, and energy map. Small, medium, and large UNet configurations were available and were trained separately. Models were optimized with AdamW using a learning rate of 1×10^-3^ and weight decay of 1×10^-4^. Training used mixed precision on GPU, gradient-norm clipping at ‖g‖_2_ ≤ 1.0, and early stopping based on the unweighted mean validation loss across output heads. The learning rate was linearly warmed up from 1% of the base rate over the first three epochs and then followed a cosine decay schedule.

The four output heads were balanced using a per-epoch weight schedule. During the first 10 epochs, losses for the cell mask, boundary, center, and energy heads were weighted as [1.0, 1.8, 3.0, 3.0]. These weights were then linearly changed over 50 epochs to [1.0, 1.3, 0.5, 0.5], keeping the cell-mask loss fixed while reducing the relative influence of the auxiliary center and energy heads.

Cell-mask and boundary losses used a weighted combination of binary cross-entropy with logits and soft Dice loss. The boundary target was taken from the dataset target channel and smoothed with a Gaussian filter (σ = 1 px) before loss calculation. The center loss combined positive-class-weighted binary cross-entropy (pos_weight = 10) and mean squared error between predicted probabilities and the target center map, both restricted to pixels inside the ground-truth cell mask. For optional center sparsity regularization, small allowed regions were generated around transformed ground-truth centers, but the sparsity and count-matching terms were set to zero in the final training setting. The energy head regressed the normalized energy target inside cells using a weighted sum of L1 and mean squared error on the sigmoid output.

The total training loss was the weighted sum of the four head losses plus a boundary-exclusion term and halo regularizers. The exclusion loss penalized high cell probability at pixels with high boundary target values. Halo regularization penalized cell-mask leakage in a narrow outer ring around cells and penalized center and energy leakage in both this ring and distant background. Model selection and early stopping were based only on the unweighted mean of the per-head validation losses. Models were trained for a maximum of 250 epochs. Final model selection was based on visual accuracy of real microscopy images and images taken by smartphones.

### Image segmentation and cell identification

For inference, we apply a UNet to full-resolution images using an internal tiling scheme. The input image is split into overlapping tiles of fixed size, with constant zero-padding at the borders if needed. Tiles are processed in batches by the network to obtain per-pixel class probabilities, which are then stitched back to full-image probability maps by averaging in the overlap regions. From the resulting cell probability map we build a robust foreground mask using a two-threshold hysteresis procedure, followed by standard morphological operations to close gaps, fill holes, and remove small objects. Instance segmentation is then formulated as a marker-controlled watershed on an “elevation” image that combines several cues: normalized distance transform inside the cell mask (favoring object centers), boundary probabilities, gradients of the cell probability map, and an optional energy map. Seed markers are obtained mainly from the center probability map using non-maximum suppression (or simple thresholding), with a fallback to connected components of the mask when no seeds are found. This probability-driven watershed separates touching objects that would otherwise be merged by a simple connected-component labeling, and allows us to control splitting and noise suppression through a small set of interpretable parameters. Full implementation details are given in the provided source code.

### Non-convolutional image segmentation pipeline (NCISP) and human cross-annotation

Images were processed with a custom ImageJ (1.54p^29^) macro that combined automated segmentation with manual annotation and was specifically designed around optimal segmentation of the color images acquired at the Olympus CKX53 microscope setup. Red, green, and blue channels were split, and the red and green channels were summed, converted to 8-bit, corrected for background (rolling ball background subtraction radius 20 and sliding paraboloid enabled combined with a flat background subtraction value 40), thresholded with the Phansalkar local thresholding method (radius = 2), and separated by standard Fiji watershed. Objects were detected with *Analyze Particles* using a size filter of 10-50 px and a circularity filter of 0.80-1.00. For each segmented region, the macro measured mean red, green, and blue intensities from read-only channel copies and assigned a unique ROI ID. In a version for human annotation, the macro subsequently displayed a merged RGB image and paused twice so that human expert annotators could mark all positive and negative cells with the multi-point tool. Each marked point was sampled for its RGB values and saved with its own ROI ID and label (user_green or user_red). All segmented ROIs and all user-selected points were written to a single CSV file containing folder name, image name, ROI ID, ROI type, and RGB means. Full technical details, including the mapping-table reader, resume logic, and window-handling steps, are provided in the source code. In total, 420 images, equally divided amongst four annotators total, were used that were also previously scored by two independent annotators as part of a routine diagnostic procedure.

### Human PRA scoring

A total of 38 assays were scored independently by two annotators with experience in routine histocompatibility diagnostics. Each annotator estimated the fraction of positive cells based on the image alone and assigned a score using fixed cutoffs: 1 for ≤10%, 2 for ≤20%, 4 for ≤50%, 6 for ≤80%, and 8 for higher values. The two sets of scores were compared to assess agreement, and both were used in later analyses.

### Automated population detection in flow cytometry

Flow cytometry events were read in using the flowio library (https://github.com/whitews/FlowIO) into a custom file representation and first subjected to quality control. Edge events were excluded based on the configured scatter channels. Where FSC-A and FSC-H were available, singlets were identified using a median absolute deviation-based gate applied to the relationship between FSC-A and FSC-H. Marker channels were transformed using an inverse hyperbolic sine transformation. Marker-specific transformation cofactors and positivity thresholds were estimated globally from quality-controlled events across the dataset using two-component Gaussian mixture models.

Lymphocyte gating was performed in FSC/SSC space using a marker-informed back-gating strategy. Events positive for any configured lineage marker were first identified using the learned marker thresholds. The union of these marker-positive events was used as a guide population for identifying the lymphocyte region in scatter space. A two-dimensional Gaussian mixture model was fitted to FSC/SSC values of quality-controlled events, and mixture components contributing to the marker-positive guide population were selected. Events assigned to these components with sufficient posterior probability were retained, followed by application of final scatter-based limits to reduce inclusion of low-FSC and high-SSC events.

Population detection was performed within the lymphocyte gate. For each event, the feature representation consisted of log-transformed FSC and SSC values together with asinh-transformed marker intensities. A pooled training set was generated across files using random per-file subsampling, with additional enrichment for events from high marker-expression tails to improve detection of less frequent marker-positive populations. Features were scaled using a median absolute deviation-based scaler and clustered using the configured unsupervised clustering algorithm.

Clusters were assigned to population labels based on their marker-expression profiles. For each cluster and marker, the fraction of events above the learned marker threshold and the median transformed marker intensity were calculated. Clusters with low median FSC and SSC values were classified as debris. Remaining clusters were assigned to the marker with the strongest evidence of positivity, provided the fraction-positive and median-intensity criteria were met. Clusters without a qualifying marker, or clusters with ambiguous marker assignment, were labeled as unknown. Marker-defined populations were then generated by assigning events according to the label of their predicted cluster.

IgG binding was quantified for all output gates, including all edge-filtered cells, singlets where available, lymphocytes, and each marker-defined population. IgG values were extracted from the configured IgG channel and analyzed both in raw scale and after inverse hyperbolic sine transformation. Gate-specific IgG positivity cutoffs were estimated from negative control samples; where positive control samples were available, these were included in cutoff estimation and fluorescence-index calculation. For each gate, the reported readouts included event count, fraction of IgG-positive events, raw and transformed median IgG signal, median shift relative to the negative control, median ratio relative to the negative control, and fluorescence index relative to the negative and positive controls. Per-file readouts were combined across files to obtain sample-level results.

## Results

AlloViewer was developed as a workflow tool for routine cell-based HLA antibody diagnostics rather than for a single standardized experimental setup. We therefore first defined the technical variability that an automated interpretation pipeline would need to tolerate in practice. In complement-dependent cytotoxicity (CDC)-based assays, microscopy images may differ in camera system, acquisition settings, illumination, staining quality, background fluorescence, cell density, cell clustering, and the proportion of viable and lysed cells. Flow cytometry crossmatch (FCXM) data are similarly variable, with experiment-specific differences in event density, scatter profiles, population geometry, and immunoglobulin G (IgG) fluorescence distributions. When developing a pipeline for automated interpretation of diagnostic assays, the pipeline therefore needs to handle heterogeneous laboratory outputs, including differences between experiments within one laboratory and differences between laboratories.

### Simulation-based image generation captures heterogeneous CDC assay and acquisition conditions

We chose a deep-learning-based approach for cell detection in CDC microscopy images because a conventional rule-based image analysis pipeline would be unlikely to generalise well across the range of camera systems, acquisition settings, staining patterns, and assay conditions expected in routine use (**Figure 1A**). However, supervised neural network training requires large numbers of representative examples. Acquiring and manually annotating such images, with potentially thousands of cells per well that would have to be masked manually, would have been labour-intensive and would still not have covered the full spectrum of potential image appearances. We therefore implemented a simulation-based strategy that allowed us to generate large numbers of CDC-like images while systematically introducing controlled variability. This approach not only gives access to the required number of images as well as the required variability but also naturally comes with a full and perfectly accurate set of annotations for each individual simulated cell.

**Figure 1.**
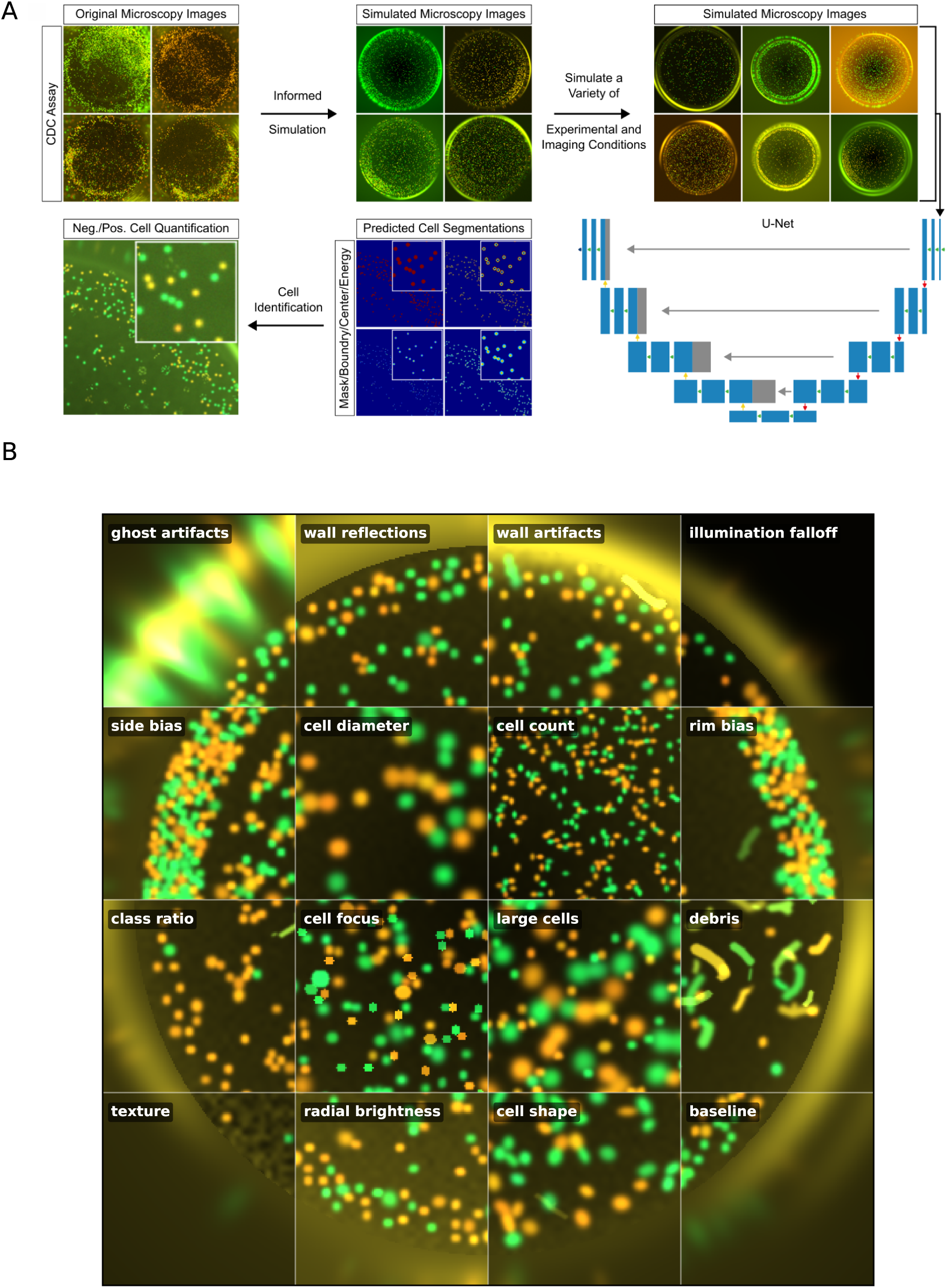
Simulated complement-dependent cytotoxicity microscopy images provide annotated training data for UNet-based lymphocyte segmentation. **A** Schematic overview of the simulated image generation, UNet model training, and subsequent UNet-based lymphocyte segmentation workflow. Synthetic fluorescence microscopy images with known cell numbers and segmentation masks were generated to train and evaluate the model under controlled and variable imaging conditions. **B** Simulation showcase illustrating the range of image features generated by the simulator. Each tile is taken from the same spatial position in an independently simulated 512 × 512 image, preserving the global well geometry while highlighting one varied parameter group. The examples show variation in ghost artifacts, wall reflections, wall artifacts, illumination falloff, spatial cell bias, cell diameter, cell density, rim bias, cell class ratio, focus, large cells, debris, background texture, radial brightness, cell shape, and the baseline simulation. *Ghost artifacts* are blurred out-of-focus cell-like signals outside the well; *wall reflections* and *wall artifacts* are supposed to depict reflections near the well boundary; *illumination falloff, radial brightness,* and *texture* describe spatial background variation. *Side bias* and *rim bias* describe non-uniform cell placement, *class ratio*, the relative abundance of green and orange cells, and *focus*, the sharpness of the cell signal. Ghosts, debris, reflections, and background artifacts were excluded from the ground-truth masks, so the network was trained to segment true in-well lymphocyte objects rather than artifact structures. For the full set of varied parameter groups refer to the **Extended Data Figures**. Neg., negative; Pos., positive.

Specifically, we created a dedicated image-simulation function to generate realistic fluorescence microscopy images resembling routine CDC assay wells across different imaging conditions, cell densities, staining qualities, cell clustering patterns, and background fluorescence intensities, among others (**Figure 1B, Supplementary Figure S1, Extended Data Figures 1-6, Methods**). Image resolutions and dimensions were simulated to resemble high-resolution modern images (up to 2560 px in width) in different aspect ratios. First exploratory experiments were promising and showed high resemblance of the simulated images to images obtained from real-world assays. However, we quickly noticed that histograms of specialized high-quality color and monochrome digital microscope cameras, as well as more accessible imaging systems such as smartphone cameras, produced markedly different histograms when imaging the same wells (**Figure 2A**). To reproduce this variability across laboratory imaging systems, we created specialized functions that would adjust the simulated histograms to reflect the diverse range found in real-world images, thereby substantially improving segmentation accuracy (**Figure 2**).

**Figure 2.**
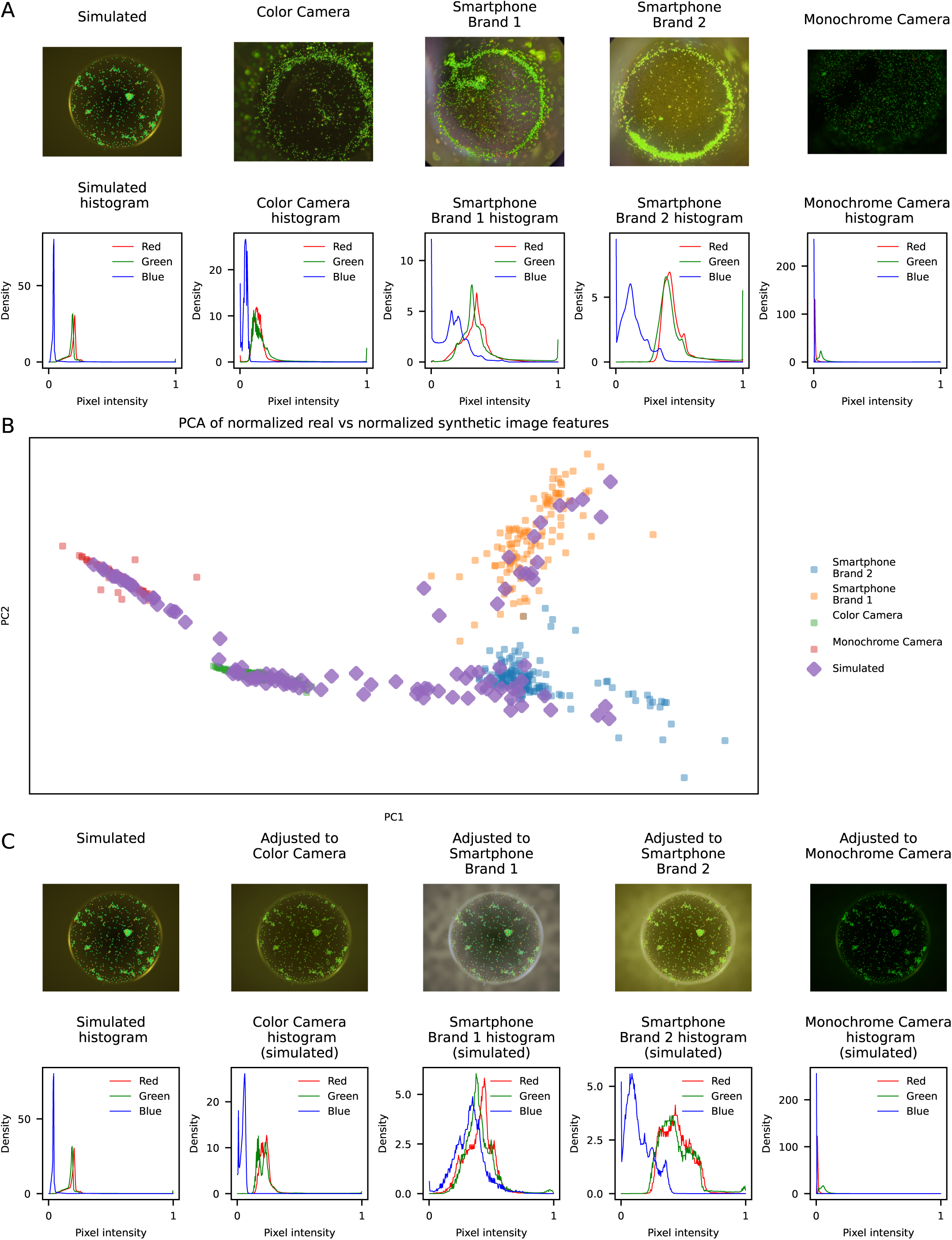
Camera-style modeling adapts simulated complement-dependent cytotoxicity microscopy images to real microscope and smartphone acquisition domains. **A** Example images from the simulated pipeline and three real acquisition domains (Microscope equipped with color and monochrome digital cameras, smartphone of brand 1, smartphone of brand 2), together with per-channel pixel-intensity histograms for the red, green, and blue channels. The real domains show clear differences in color balance, background structure, and intensity distribution. **B** Principal component analysis (PCA) of normalized image features extracted from real images (n=649) and normalized synthetic images (n=400). Real images from the four acquisition domains form distinct clusters, and the simulated images partially overlap with the real domains after style adjustment (plotted are 100 images per domain for better visibility). Histogram adherence has been set to high values. The images in the training dataset were generated with lower adherence values. **C** Example simulated image after domain-specific camera-style adjustment to match the appearance of each real acquisition domain, shown together with the corresponding per-channel histograms. After adjustment, the simulated images show domain-dependent changes in color composition, contrast, and intensity distribution that more closely resemble the target real-image domains.

Ultimately, we created a comprehensive training dataset of 50.000 images that covered a wide variety of different image simulation parameters equally (**Supplementary Figure S2**). In order to ascertain a confident cell segmentation, we aimed to define cells by their area, their outlines, their center localization and the relative center distance to edges^30,31^ and co-simulated these images as later target images for segmentation (**Supplementary Figure S3**).

### Full-resolution tiling improves UNet-based lymphocyte segmentation

A convolutional neural network (CNN) with the UNet-architecture was chosen for the segmentation task as UNet-based models are especially suited for pixel-level segmentation of biomedical images^17^. To develop a transferable approach that could also be deployed through external web-based services as well as locally on non-specialized hardware, we trained UNet models of different sizes to identify the smallest architecture that still provided adequate segmentation performance (see **Methods**). Since the input image dimensions into the CNN are quadratic and were chosen to be 512 × 512 pixels, images had to be preprocessed in order to conform with these requirements. We therefore compared three preprocessing strategies. In the first approach, images were zero-padded to obtain a square format and then resized to the target dimensions (“pad_resize”). In the second approach, a local thresholding method was used to approximate the well outline, followed by well-based cropping and resizing (“crop_well_resize”). In the third approach, images were divided into partially overlapping 512 × 512 pixel tiles to preserve the original image resolution while reducing edge artifacts during downstream reconstruction (“tiles”; **Supplementary Figure S1**).

During training, all models showed a rapid and stable reduction in loss values (**Supplementary Figure S4**). However, models trained on tiled images achieved substantially lower validation loss values than models trained on padded or cropped images (**Supplementary Figure S4A**). This difference was partly attributable to the loss of single-cell resolution during resizing of padded or cropped full-well images, where individual lymphocytes were frequently reduced to diameters of only 1-3 pixels or disappeared entirely, resulting in impaired segmentation accuracy (data not shown). Consistent with this observation, mask-level and cell-center detection accuracy depended strongly on the preprocessing strategy, whereas UNet model size had a smaller effect (**Supplementary Figure S4 and S5**). We therefore selected full-resolution tiled images for all subsequent segmentation analyses. To finally convert the resulting model predictions into individual cell objects, we implemented a custom instance-segmentation workflow based on watershed seeding and probability maps (**Figure 3A and B**).

**Figure 3.**
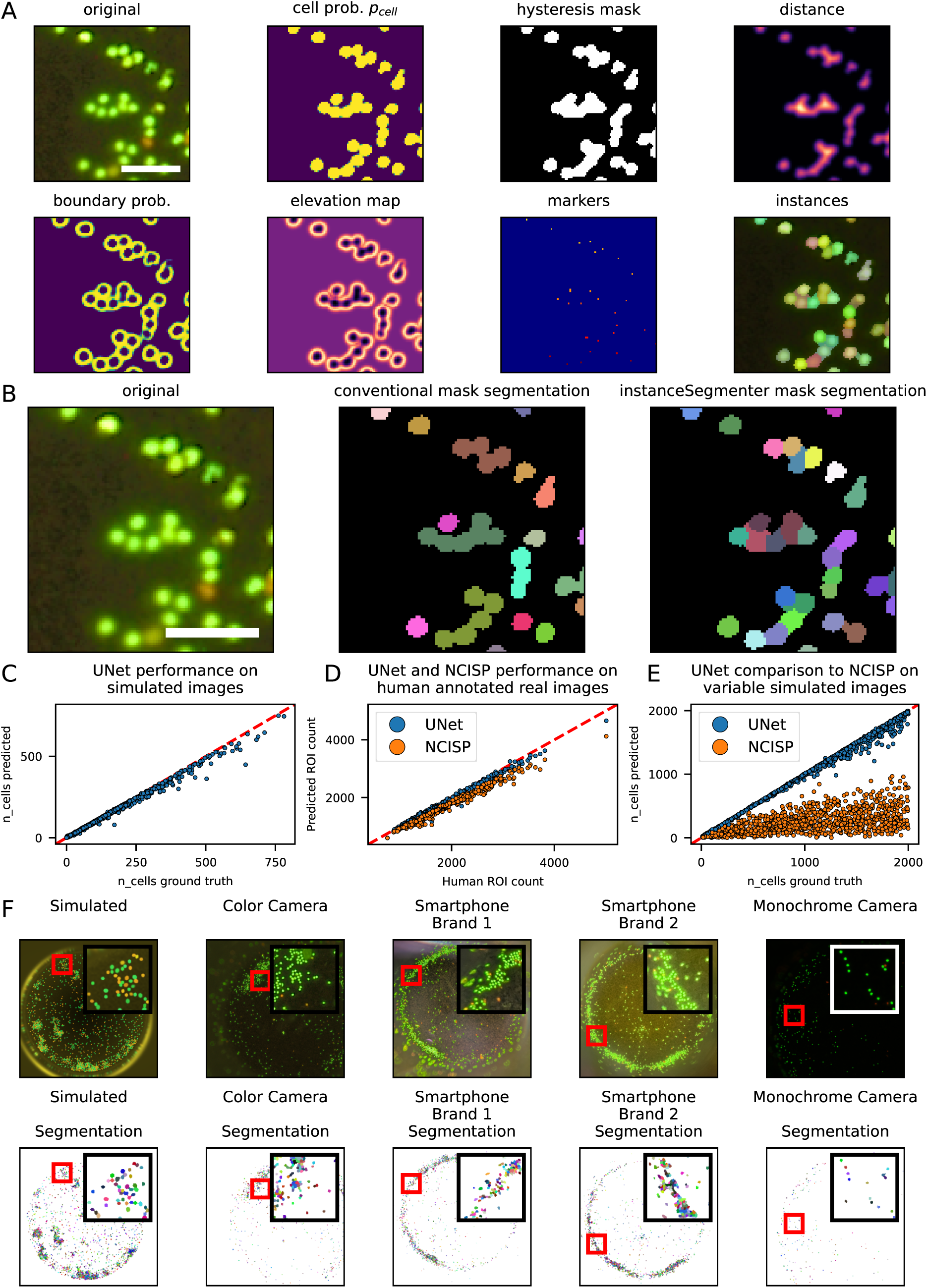
Development, validation and generalization of the UNet-based instance-segmentation workflow. **A** Step-by-step visualization of the custom instance segmentation procedure for one cropped tile. The raw fluorescence image is passed through the UNet to obtain cell mask, boundary, center probability maps as well as a density estimate. The cell probability map is converted into a stable binary mask using hysteresis thresholding, followed by distance-map calculation. Boundary and distance information are combined into an elevation map, while center probabilities are used to define watershed markers. The final panel shows the resulting labeled cell instances. **B** Comparison of two segmentation strategies on the same cropped region. Conventional mask segmentation uses thresholding and connected-component labeling, while the InstanceSegmenter uses the full watershed-based pipeline shown in panel A. The InstanceSegmenter better separates touching cells and produces cleaner instance labels. **C** Quantitative validation of UNet-based cell counting on an independent simulated test dataset. Each point represents one image tile (n=2000). The x-axis shows the ground truth number of simulated cells, and the y-axis shows the number of cells detected by the UNet-based instance-segmentation workflow. The dashed line indicates perfect agreement between ground truth and prediction. **D** Comparison of UNet-based and rule-based cell counting with manual human annotations in experimentally acquired real microscopy images. The rule-based comparator was implemented as a custom non-convolutional image segmentation pipeline (NCISP) in ImageJ. Each point represents one manually annotated image (n=420). The x-axis shows the human-annotated cell count, and the y-axis shows the corresponding algorithm-derived cell count. The dashed line indicates perfect agreement. UNet-derived cell counts showed near-perfect agreement with manual annotations, whereas NCISP performance decreased at higher cell densities and in clustered images. **E** Comparison of UNet-based and NCISP-based cell counting on simulated images with variable imaging and assay conditions. Each point represents one simulated image (n=1000). The x-axis shows the ground truth number of simulated cells, and the y-axis shows the predicted cell count. Points are colored according to the analysis method. The dashed line indicates perfect agreement with ground truth. In contrast to the UNet-based workflow, the rule-based NCISP did not reliably generalize to heterogeneous simulated image conditions. **F** Representative input images and corresponding UNet-based instance-segmentation outputs. The upper row shows cropped input images from five image sources: simulated images, microscope-camera images of two camera types, and smartphone images of two different brands. The lower row shows the corresponding predicted instance segmentations. Individual segmented objects are displayed in different colors, and background pixels are shown in black. Red boxes mark enlarged regions shown as insets in the upper-right corner of each panel to illustrate local segmentation quality. Together, these analyses show that the simulation-trained UNet workflow accurately recovers cell counts from simulated images with known ground truth, agrees with human annotations in experimentally acquired real microscopy images, and remains applicable to images acquired with different imaging devices under differing imaging conditions.

### Deep learning-based segmentation outperforms rule-based image analysis in heterogeneous CDC images

We next evaluated the performance and generalizability of the final segmentation workflow. Upon manual visualization of the models ability to segment cells, we observed a noticeable increase of model-accuracy during the first epochs, regardless of image origin. Crucially, later models often failed to detect cells reliably in images that were derived from real cameras. We treated this as a classical overfit, where a model, trained on simulated images, would at some point be too familiar with this specific type of cells while not being able to generalize across modalities. Consequently, validation was performed with a model that visually performed well on all acquired image types.

For evaluation we first tested the UNet-based pipeline, including custom instance segmentation, on an independent set of simulated CDC images that was not trained on. This confirmed high agreement between predicted and expected cell counts, as well as robust performance for cell-center detection and mask generation (**Figure 3C and Supplementary Figure S6**). To benchmark this approach against conventional image analysis, we developed an independent rule-based, non-convolutional image segmentation pipeline (NCISP) using classical image processing operations. This pipeline was manually optimized on real microscopy images acquired under routine assay conditions (see **Methods**) and achieved acceptable performance on selected representative images. However, despite extensive optimization, the NCISP did not reliably resolve dense cell clusters (data not shown). We therefore used human-annotated real images as an additional reference standard for performance comparison on real microscopy data (see also **Methods**). Cell counts derived from the UNet-based workflow showed near-perfect concordance with human annotations, whereas the NCISP became increasingly inaccurate at higher cell densities (**Figure 3D**). When applied to simulated images designed to reflect broader inter-assay and inter-laboratory variability, the NCISP failed to segment cells reliably across the generated image conditions (**Figure 3E and Supplementary Figure S7**). These findings indicate that variability in assay and imaging conditions can readily exceed the robustness of a fixed rule-based image analysis pipeline, whereas the simulation-trained UNet approach remained suitable for heterogeneous image inputs. Consistent with this, real images acquired with different microscopy camera types as well as smartphone cameras were also segmented successfully by the UNet-based workflow (**Figure 3F**).

### Controls-calibrated Gaussian classification enables automated CDC cytotoxicity scoring

In CDC assays, cytotoxicity is assessed by differential fluorescence staining of viable and lysed cells. Lysed cells take up the red fluorescent dye and appear orange to red, whereas viable cells remain predominantly green. After UNet-based segmentation, each detected cell was represented as a region of interest (ROI), from which red and green fluorescence intensities were extracted. To classify individual ROIs in an experiment-specific manner, AlloViewer used the internal positive and negative control wells included in each assay. ROIs from positive-control wells were used to define the fluorescence distribution of lysed cells, whereas ROIs from negative-control wells were used to define the fluorescence distribution of viable cells (**Figure 4A**).

**Figure 4.**
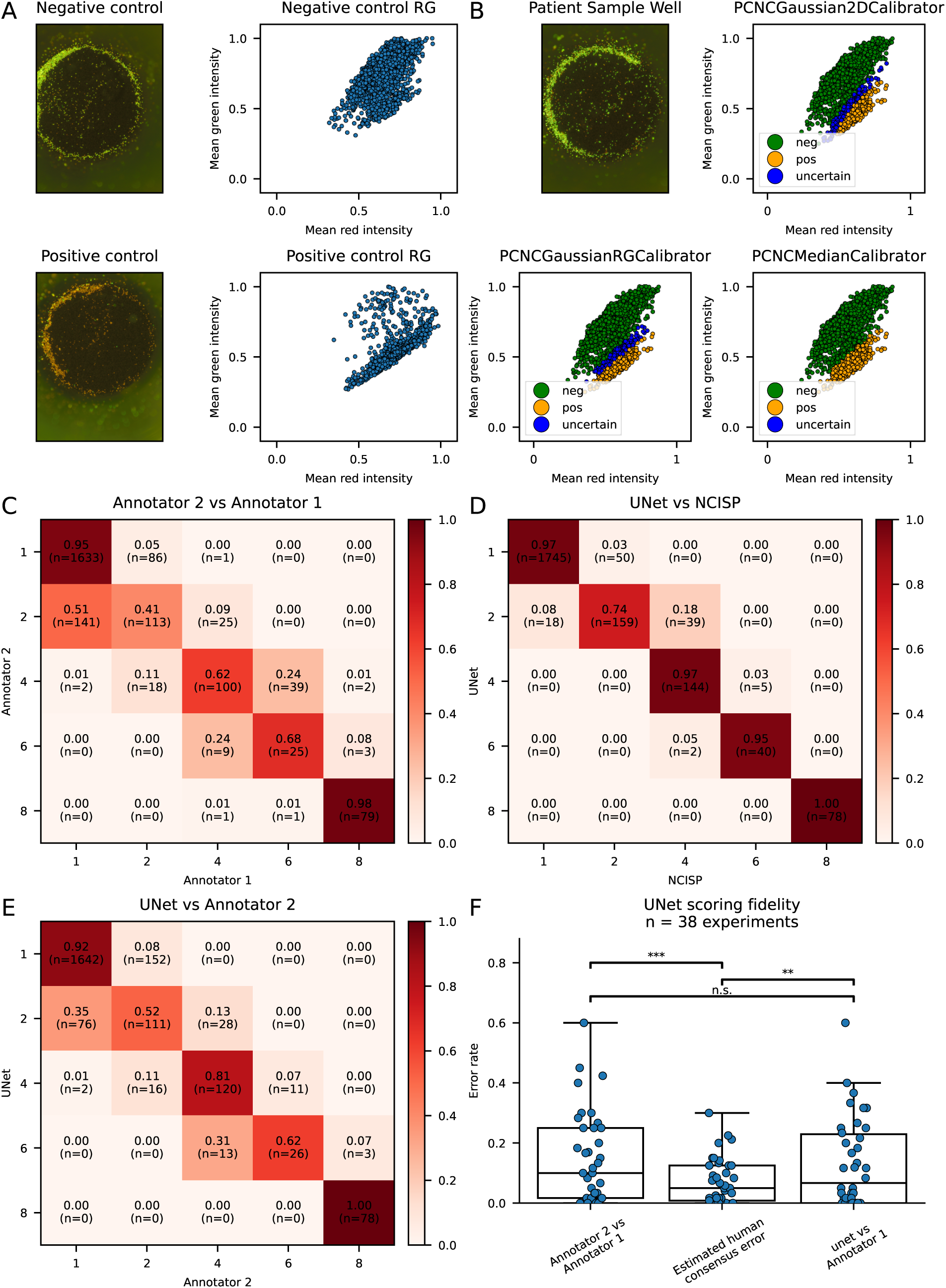
Control-calibrated cell classification and scoring fidelity of the automated complement-dependent cytotoxicity workflow. **A** Representative negative- and positive-control wells with corresponding red-green fluorescence intensity scatter plots of segmented cells. Negative-control wells show a predominantly green signal distribution, whereas positive-control wells show a shift toward increased red fluorescence. **B** Representative patient sample well with object-level red-green intensity classification using three calibration strategies. Segmented objects were classified as negative, positive, or uncertain according to their red and green fluorescence intensity profiles. Object colors indicate negative cells in green, positive cells in orange, and uncertain cells in blue. **C** Confusion matrix comparing scores assigned on real images by Annotator 2 with scores assigned by Annotator 1. Rows indicate Annotator 2 scores, and columns indicate Annotator 1 scores. Values show row-normalized fractions, with raw counts shown in parentheses. The matrix illustrates the degree of inter-annotator variability in manual complement-dependent cytotoxicity scoring (n=2280 images from n=38 experiments). **D** Confusion matrix comparing well scores obtained after UNet-based segmentation with scores obtained after ImageJ-based non-convolutional image segmentation pipeline (NCISP) segmentation, while keeping the downstream classification and scoring procedure identical. The strong overlap in agreement indicates that final categorical well scores were largely stable across segmentation approaches on real images with imaging conditions for which the NCISP was specifically designed for (n=2280 images from n=38 experiments). **E** Confusion matrix comparing automated UNet-based well scores with Annotator 2 scores on real images (n=2280 images from n=38 experiments). **F** Experiment-level scoring fidelity of the automated UNet-based workflow. Each point represents one experiment. The left box shows the inter-annotator error rate between Annotator 2 and Annotator 1. The middle box shows the estimated human-consensus error, calculated as half of the inter-annotator error rate. The right box shows the automated UNet-based error rate relative to Annotator 1. The automated workflow showed a numerically lower error rate than Annotator 2 when both were compared with Annotator 1, although this difference was not statistically significant by paired Wilcoxon signed-rank test. RG, Red-Green; neg, negative; pos, positive. *** p<0.001, n.s. not significant.

We compared several control-based calibration strategies for ROI classification. These included simple thresholding of the red-to-green intensity ratio using positive- and negative-control medians or means, a one-dimensional Gaussian model of the red-to-green ratio, and a two-dimensional Gaussian model based on the joint red and green intensity space. The final classifier used the two-dimensional Gaussian approach. For each experiment, separate bivariate normal distributions were fitted to ROIs from the positive- and negative-control wells. Each sample ROI was then assigned a classification score based on the difference between its log-likelihood under the positive-control and negative-control distributions. ROIs were classified as positive, negative, or uncertain depending on whether this log-likelihood difference exceeded a predefined margin (**Figure 4B**).

The two-dimensional Gaussian classifier was selected for downstream analysis because it preserved the joint structure of the red and green fluorescence signals instead of reducing each ROI to a single red-to-green ratio. This was important because positive and negative ROI populations were not separated solely by a simple shift in the red-to-green ratio. Rather, their spread and covariance in red-green intensity space also contributed to the classification boundary. The classifier therefore allowed ROIs located near the overlap between positive- and negative-control distributions to be marked as uncertain instead of forcing a binary decision.

We next assessed automated scoring fidelity by comparing AlloViewer-derived well scores with scores from two independent human annotators. Since no objective biological ground truth exists for microscopic CDC scoring, automated performance was benchmarked against the practical reproducibility limit imposed by human inter-observer variability. Following the logic of the human-consensus bound^31^, disagreement between Annotator 1 and Annotator 2 was used to estimate human-level variability, and half of this disagreement rate was taken as the expected error relative to an unobserved human-consensus score (**Figure 4C**).

To determine whether the final well-level readout depended on the segmentation method, we also compared NCISP-based and UNet-based segmentation while keeping the downstream classification and scoring procedure identical (**Figure 4D**). The strong diagonal structure of the resulting confusion matrix indicates that categorical well scores were largely stable across segmentation approaches. Note that the NCISP was designed for the imaging conditions of the real images used in this comparison specifically and, as shown in Figure 3E, is not applicable outside of these.

Finally, we compared experiment-level scoring errors between the automated workflow and human annotation. When both were compared against Annotator 1, the UNet-based workflow showed a numerically lower experiment-level error rate than Annotator 2 (**Figure 4F**). Its error rate was slightly higher than the estimated human-consensus error, which represents the theoretical lower bound derived from half of the inter-annotator disagreement rate. These results therefore indicate that AlloViewer achieved scoring performance within the range of human inter-annotator variability.

### Marker-informed clustering enables automated population identification in flow cytometry crossmatch data

We next extended AlloViewer from image-based CDC analysis to flow cytometry crossmatch (FCXM) interpretation as it is the other diagnostic option of pre-transplant crossmatching. In contrast to CDC microscopy, FCXM provides quantitative fluorescence readouts at the single-event level, but post-acquisition analysis still depends on manual gating in specialized software and expert interpretation of population-specific immunoglobulin G (IgG) signals. We therefore developed an automated, unsupervised but marker-informed workflow for identifying relevant cell populations in FCXM data (see **Methods**). After quality control and lymphocyte gating, events were clustered in transformed feature space and assigned to marker-defined populations using calibrated marker thresholds. Population-specific IgG metrics were then calculated using gate-specific cutoffs derived from the control samples.

To identify a suitable clustering strategy, we benchmarked representative graph-based^32^, self-organizing map-based^33^, and density-based methods^34,35^ across a range of parameter settings. This comparison was designed to identify an approach that provided reliable population detection while maintaining moderate computational requirements for routine use (**Supplementary Figures S8 and S9**). Based on this benchmark, we selected the final algorithm and parameter settings used for the main FCXM analysis shown in **Figure 5**. Across the evaluated populations in the dataset, the workflow achieved high F1 scores, indicating accurate automated identification of marker-defined subpopulations. In order to evaluate if the anti-IgG signal actually corresponds to the presence of a meaningful binding of anti-IgG-antibodies, laboratories have established laboratory-specific metrics, among them the difference of medians, the fold-change of medians or the percentage of IgG-positive cells. Here, we implemented all these metrics to comply with those laboratory-specific standards.

**Figure 5.**
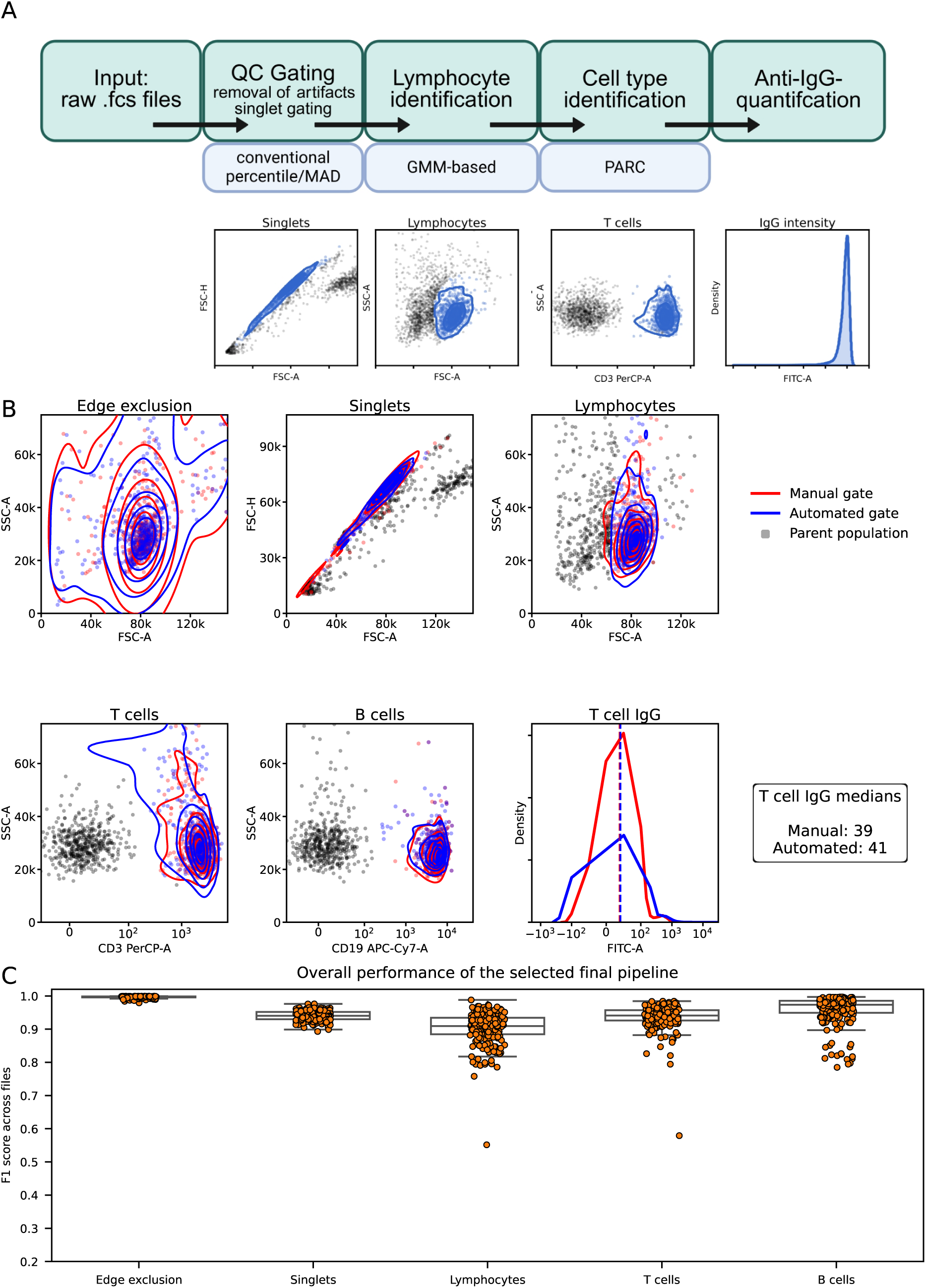
Automated flow cytometry crossmatch analysis closely matches manual gating across the analysis workflow. **A** Schematic overview of the automated flow cytometry crossmatch (FCXM) analysis pipeline. Raw .fcs files first undergo quality-control gating to remove artifacts and select singlet events, followed by lymphocyte identification, marker-informed cell-type assignment, and population-specific anti-immunoglobulin G (IgG) quantification. Representative plots illustrate the successive intermediate populations and the final IgG intensity readout. **B** Representative sample-level comparison of manual and automated gating across the analysis workflow. For each gating step, parent events are shown in gray, manually gated events in red, and automatically gated events in blue. Contour lines indicate event-density estimates (up until the 5th percentile). The final panel shows the IgG intensity distributions of manually and automatically gated T cells, together with the corresponding median IgG values. **C** Overall performance of the selected final pipeline across analyzed files (223 files of 23 independent experiments). Boxplots with individual sample-level points show F1 scores for each gating target, including edge exclusion, singlet selection, lymphocyte identification, T-cell identification, and B-cell identification. Automated gating showed high agreement with manual gating across files, with strongest performance for upstream quality-control gates and robust performance for lymphocyte and downstream cell-type identification. QC, quality control; MAD, median absolute deviation; GMM, gaussian mixture model.

### Web-based AlloViewer implementation supports CDC test-cell panel analysis, CDC crossmatch, and FCXM interpretation

To make the automated analysis workflow accessible across assay formats and laboratory settings, we implemented AlloViewer as a web-based platform with a Python backend and a TypeScript/React frontend. The interface guides users through assay-specific analysis modes beginning with selection of the experimental layout. Three diagnostic workflows are currently supported: CDC-based HLA antibody specificity analysis using commercial test-cell panels, CDC crossmatch analysis, and FCXM analysis (**Figure 6**).

**Figure 6.**
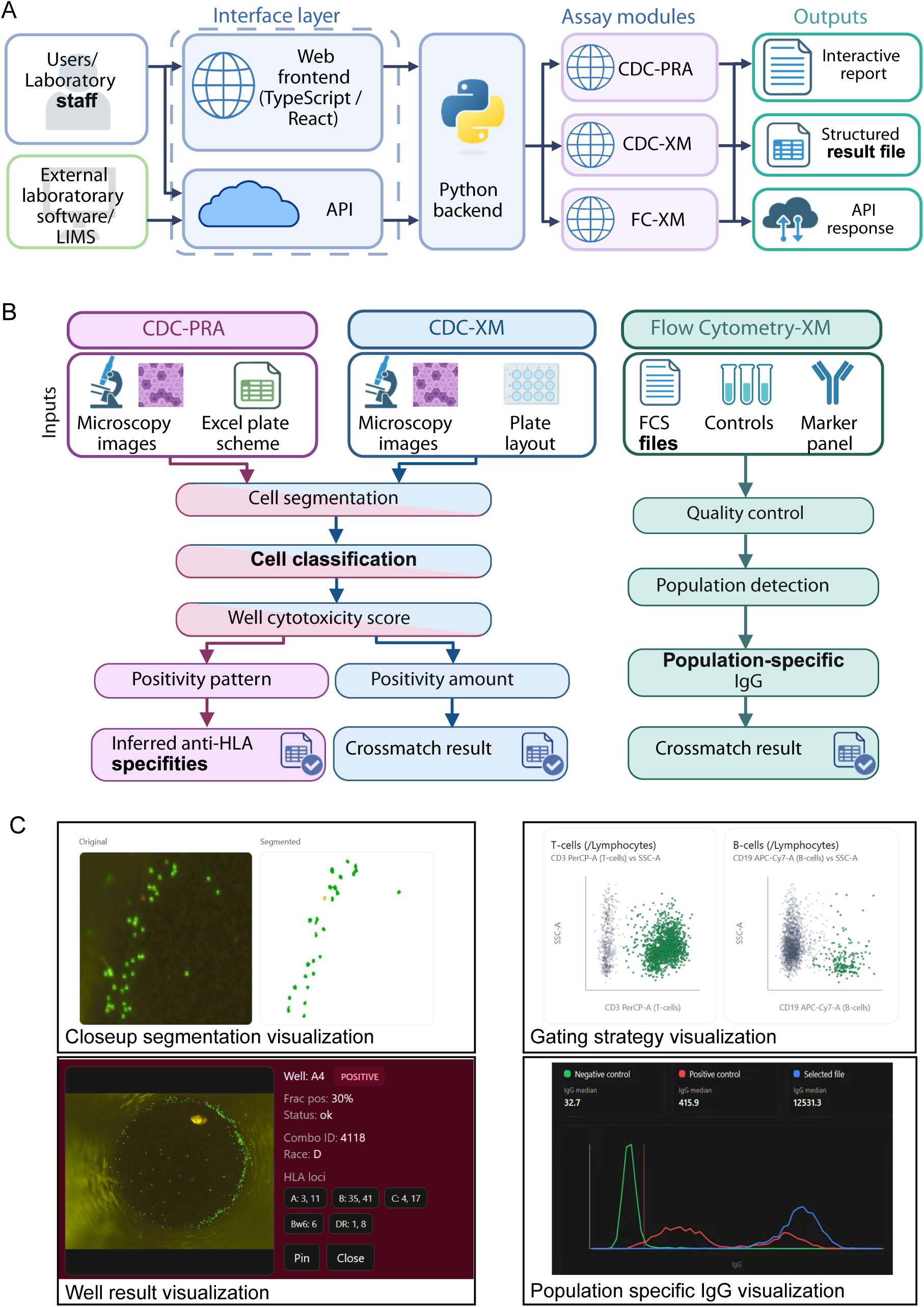
AlloViewer platform architecture and API-enabled diagnostic workflow. **A** AlloViewer was implemented as a web-based analysis platform with a TypeScript/React frontend, a Python backend, and an application programming interface (API). Users and laboratory staff can access the assay modules through the graphical web interface, while external laboratory software or laboratory information management systems (LIMS) can submit data and retrieve results through the API. The same backend analysis modules are used for CDC-PRA, CDC-XM, and FCXM, producing interactive reports, structured result files, or API responses. **B** Summary of the three implemented assay workflows. For CDC-PRA, microscopy images are combined with an uploaded Excel plate scheme, followed by cell segmentation, cell classification, well-level cytotoxicity scoring, and inference of likely anti-HLA specificities from the resulting positivity pattern. For CDC-XM, microscopy images and the crossmatch layout are processed by the same image-analysis workflow to generate a well-level cytotoxicity score, positivity amount, and final crossmatch result. For flow-cytometry crossmatch, FCS files, controls, and the marker panel are processed by automated quality control and population detection, followed by population-specific IgG quantification and crossmatch interpretation. **C** Representative screenshots from the AlloViewer interface showing intermediate and final analysis outputs. The image-analysis modules provide close-up segmentation views and well-level result summaries, allowing review of segmentation quality, cell-level calls, cytotoxicity scores, control status, donor/recipient metadata, and inferred HLA patterns. The flow-cytometry module provides gating visualizations and population-specific IgG plots, including comparison of the selected sample with negative and positive controls. These views allow users to inspect the steps leading to the final assay-level result rather than relying only on the final report.

For CDC-based HLA antibody specificity analysis (CDC-PRA), the platform applies the same image-based UNet segmentation and control-calibrated cell-classification workflow to commercial CDC test-cell panels used for determining the HLA antibody specificities in a patient’s serum. In this assay format, each well contains defined test cells with known HLA typing information, and antibody specificities are inferred from the pattern of positive and negative cytotoxicity reactions across the panel. The platform therefore allows upload of an standardized Excel-based panel scheme describing the HLA typing associated with each well. After automated well-level scoring, AlloViewer maps the observed reactivity pattern to the uploaded test-cell panel layout and derives candidate HLA antibody specificities compatible with the detected cytotoxicity pattern. This enables interpretation of CDC-based test-cell panels directly from the experimental plate layout and reduces the need for manual comparison of reactive wells against the panel scheme.

For CDC crossmatch assays, AlloViewer applies the image-based segmentation and control-calibrated cell-classification workflow described above to wells containing donor lymphocytes incubated with recipient serum. The software detects individual cells, classifies cell-level viability staining, calculates well-level cytotoxicity fractions, and reports the resulting crossmatch interpretation according to the selected scoring rules.

For FCXM assays, uploaded flow cytometry standard .fcs files are processed through the automated population-identification workflow, followed by calculation of population-specific IgG readouts and assignment of the final crossmatch result. The same web interface therefore supports both image-based and flow-cytometry-based crossmatch interpretation while preserving assay-specific preprocessing, classification, and scoring logic.

In addition to the graphical user interface, AlloViewer provides a documented application programming interface (API), allowing external systems to submit assay data, retrieve processed results, and integrate the analysis workflow into existing laboratory software environments. This architecture separates user interaction, computational analysis, and programmatic access, enabling both manual web-based use and automated integration into higher-throughput diagnostic workflows. Together, the implementation translates the segmentation, cell-classification, and flow-cytometry analysis components into an accessible software environment for standardized interpretation of heterogeneous histocompatibility assay outputs.

## Discussion

### Integrated automation of cell-based HLA antibody diagnostic workflows

In this study, we developed AlloViewer, a web-based software platform for automated interpretation of cell-based HLA antibody diagnostics across image-based and flow-cytometry-based assay formats. The central aim was not only to automate a single analytical step, but to create a traceable workflow that can process heterogeneous laboratory outputs and convert them into interpreted diagnostic readouts. To this end, we combined simulation-trained UNet segmentation, controls-calibrated cell classification, automated flow cytometry population identification, a web-based implementation and an application programming interface (API). Together, these components support automated analysis of CDC-based HLA antibody specificity testing using commercial test-cell panels, CDC crossmatch assays, and flow cytometry crossmatch assays and thus address the post-analytical bottleneck of operator-dependence and subjectivity in interpretation of these diagnostics in a single software environment.

### Image simulation supports training across technical assay variation

One of the principal methodological advances of this work is the use of simulation-trained deep learning to overcome one of the central limitations of artificial intelligence (AI) development for rare or diverse diagnostic workflows: the absence of sufficiently diverse annotated training data. In diagnostic image analysis, the limiting factor is often not only the number of annotated images, but whether the available images capture the full range of technical variation encountered in routine practice. This is particularly relevant for CDC microscopy, where images may differ in camera system, image size, illumination, exposure, staining quality, background fluorescence, cell density, focus, and cell clustering. A manually annotated dataset from a single laboratory would likely cover only a restricted subset of this parameter space.

Simulation allowed us to generate large numbers of CDC-like images while systematically varying technical and assay-related parameters. This enabled training of a segmentation model that was not optimized for one fixed imaging setup, but exposed to a broader range of expected image appearances. The strategy is especially suitable for CDC microscopy because the relevant visual structures are comparatively well defined: individual lymphocytes, viable and lysed staining patterns, background signal, well borders, and common imaging artifacts. This distinguishes CDC images from more complex biological images, such as tissue sections, where realistic simulation of biological architecture is substantially more difficult.

The value of simulation in this setting therefore extends beyond increasing annotated dataset size. It provides a controlled way to expose the model to rare, extreme, or underrepresented imaging conditions that may not be available during early development. In this sense, simulation served as a form of technical stress testing during model training. However, simulation should not be understood as a substitute for validation on real diagnostic material. Rather, it is a complementary strategy to improve robustness before subsequent evaluation in experimentally acquired datasets.

### Specialized diagnostic segmentation over generalist image-analysis model

Generalist biomedical segmentation models, including Cellpose-like and foundation-style approaches^36–39^, provide powerful options for many cell and tissue segmentation tasks. However, the aim of AlloViewer was not only to identify cells in isolated images. The diagnostic task required a controlled, assay-specific pipeline that links segmentation to downstream color classification, internal controls calibration, well-level cytotoxicity scoring, CDC test-cell panel interpretation, and crossmatch reporting. For this purpose, a specialized model has practical advantages.

First, a task-specific model can be kept comparatively small, which facilitates deployment in a web-based diagnostic software environment. Second, it can be trained on image properties that are directly relevant to CDC microscopy rather than on broad but potentially less assay-specific image collections. Third, its outputs and failure modes can be more directly connected to the diagnostic workflow. This is important because segmentation in this context is not an end in itself; it is the first step in a chain of decisions that ultimately produces an assay-level interpretation. The performance requirements are therefore defined not only by object detection accuracy, but also by the stability and traceability of downstream diagnostic scoring.

### Deep-learning-based segmentation improves robustness across heterogeneous imaging conditions

Our results also illustrate why conventional rule-based image analysis is difficult to generalize across CDC microscopy conditions. A manually optimized non-convolutional image segmentation pipeline could be tuned to perform acceptably on selected images acquired under defined conditions within one laboratory’s setup. However, its performance decreased in dense fields, clustered cell regions, and simulated images with broader technical variability. This reflects a fundamental limitation of fixed image-processing pipelines, as they often rely on assumptions about contrast, object size, background intensity, and object separation. When these assumptions are violated, performance can deteriorate rapidly. The simulation-trained UNet workflow was more robust because it learned cell-related image features across a broad range of simulated conditions. This was particularly important for high-density images and clustered cells, where simple thresholding and watershed-based rules were insufficient to reliably separate individual objects. During training, later checkpoints showed increasing specialization toward simulated image appearances and reduced visual performance on camera-derived images, consistent with domain-specific overfitting; the final checkpoint was therefore selected based on preserved segmentation performance across simulated, microscopy-camera, and smartphone-derived images.

### Experiment-specific calibration enables transferable cell classification

A second important component of the CDC workflow is experiment-specific cell classification using internal positive and negative controls. Absolute red and green fluorescence intensities are affected by staining quality, microscope and camera settings, exposure, illumination, image acquisition parameters, and assay handling. A single global threshold would therefore be unlikely to transfer reliably across different runs, laboratories, or imaging devices.

AlloViewer addresses this by calibrating the classification model within each experiment. Positive-control wells define the fluorescence distribution of lysed cells, whereas negative-control wells define the distribution of viable cells. The final two-dimensional Gaussian classifier uses the joint red and green intensity space rather than reducing each cell to a single red-to-green ratio. This is important because positive and negative cell populations may differ not only in their ratio, but also in signal spread, covariance, and the degree of overlap between control distributions. The classifier therefore preserves more information from the fluorescence profile and applies the same statistical decision rule after adapting to each experimental run.

### Automated CDC scoring reaches human-level reproducibility

Comparison with human annotators showed that the automated workflow achieved scoring performance within the range of human inter-annotator variability. This represents a particularly meaningful benchmark because no objective ground truth exists for experimentally acquired CDC assay images. Manual scoring itself is inherently variable, particularly around borderline cytotoxicity thresholds (i.e. wells that could reasonably be assigned either a score of 2 (negative) or 4 (positive)) and in wells with heterogeneous staining or high cell densities containing up to several thousand cells. We therefore used disagreement between two independent annotators as an estimate of human-level variability and derived a human-consensus bound as a reference for the lowest practically achievable error relative to an unobserved consensus score^31^.

Against this benchmark, the automated workflow achieved a numerically lower experiment-level error rate than one human annotator when both were compared with the other annotator, and its performance closely approached the estimated human-consensus bound. These findings suggest that the automated workflow operates near the practical reproducibility limit imposed by human disagreement while additionally providing consistent quantitative intermediate outputs.

Nevertheless, human experts remain essential for identifying failed assays, rare artifacts, poor sample quality, unusual staining patterns, and cases requiring clinical or immunological context. Rather than replacing expert judgement, automated analysis can reduce repetitive manual work, apply predefined scoring rules consistently, preserve quantitative intermediate data, and enable re-analysis with fixed model and parameter versions. Together, these properties improve the reproducibility and traceability of routine diagnostic workflows.

### Automated flow cytometry interpretation extends the platform beyond CDC microscopy

Although the main technical innovation of AlloViewer lies in automated CDC image interpretation, the platform was designed to also address post-analytical variability in FCXM analysis. FCXM provides quantitative fluorescence information at the single-event level and is generally more directly numerical than microscopic CDC scoring. Nevertheless, interpretation still commonly depends on manual gating, expert-defined population selection, and extraction of population-specific IgG readouts in specialized software. These steps can introduce operator dependence and limit reproducibility, particularly when gating boundaries are ambiguous or when population distributions vary between samples.

The automated FCXM workflow therefore addresses a different but related problem which is not segmentation of microscopy images, but reproducible identification of relevant cytometry populations and standardized extraction of population-specific IgG metrics. By combining quality-control gating, lymphocyte selection, marker-informed clustering, calibrated population labeling, and gate-specific IgG readout calculation, AlloViewer extends the same standardization principle from CDC microscopy to flow cytometry data. The high F1 scores observed across analyzed populations indicate that automated population detection can closely reproduce manual gating for the tested datasets.

At the same time, the FCXM component should be interpreted as a workflow extension rather than as a fully general solution for all possible cytometry configurations. Flow cytometry analysis depends strongly on marker panel design, compensation, instrument settings, event quality, sample preparation, and the selected metric for positivity. Different laboratories may use different panels, thresholds, and interpretive conventions. Further laboratory-specific validation will therefore be required to determine how well the current algorithm generalizes across instruments, protocols, and crossmatch formats. Still, the results support the feasibility of integrating automated FCXM interpretation into the same diagnostic software environment as CDC-based assays.

### Web-based deployment enables assay-specific interpretation and laboratory integration

The web-based implementation was another essential part of the study because the intended output is not only a model prediction, but a usable diagnostic workflow. Cell-based HLA antibody diagnostics include assay formats with different data structures and interpretation rules. CDC-based HLA antibody identification testing using commercial test-cell panels requires linking a pattern of positive and negative wells to the HLA typing information associated with each panel well. CDC crossmatch analysis requires image-based cytotoxicity scoring in wells containing donor lymphocytes and recipient serum prior to organ transplantation. FCXM analysis requires automated population identification and population-specific IgG readouts from flow cytometry standard files.

By bringing these workflows into one software environment, AlloViewer supports consistent analysis while preserving assay-specific logic. The graphical interface allows users to select the relevant assay format, upload the required experimental layout or data files, and retrieve standardized outputs. The API further allows external systems to submit assay data and retrieve processed results, creating a path toward integration with existing laboratory information systems or high-throughput workflows. This separation between user interaction, computational analysis, and programmatic access is important for future diagnostic deployment, because laboratories differ in whether they need interactive review, automated batch processing, or integration into existing informatics infrastructure.

### Limitations of the study

Several limitations to the study remain. First, even simulated images cannot capture all real-world artifacts. Rare staining failures, uncommon debris, unusual lymphocyte morphology, extremely poor focus, sample degradation, atypical well appearances, and device-specific artifacts may still challenge the segmentation and classification workflow. Second, although simulation allows broad technical variation, biological and procedural variation remain separate issues. Donor cell quality, complement activity, patient serum properties, disease context, local protocols, staining reagents, and operator-dependent sample handling may all influence assay readouts and require broader validation. Third, human annotation is an imperfect reference standard. The estimated human-consensus bound is useful for contextualizing automated scoring performance, but it is still derived from disagreement between annotators rather than from an independent biological truth. Fourth, the present study evaluates analytical performance and workflow feasibility, but has yet to be evaluated within prospective multi-center validation before the system can be considered for routine clinical decision support.

The flow cytometry component also requires laboratory-specific validation. The current workflow demonstrates that automated gating and population-specific IgG extraction can reproduce manual analysis in the tested setting, but generalizability across laboratories, marker panels, acquisition protocols, and interpretive thresholds remains to be established. In addition, the optimal FCXM positivity metric may depend on local practice and clinical context. Future work should therefore compare alternative IgG readouts, assess robustness across cytometer platforms, and determine how automated FCXM results align with established laboratory interpretations and clinical outcomes.

### Conclusion

In summary, AlloViewer provides an integrated framework for automated interpretation of CDC microscopy and FCXM data for pre-transplant histocompatibility diagnostics. Simulation-trained deep learning enabled robust CDC image analysis across heterogeneous imaging conditions, while experiment-specific fluorescence calibration achieved automated cytotoxicity scoring with performance approaching the practical reproducibility limit defined by human inter-annotator agreement. Together with automated FCXM interpretation and web- and API-based deployment, AlloViewer establishes a unified computational workflow for standardized, traceable interpretation of the major cell-based HLA antibody assays. These findings demonstrate the potential of simulation-trained AI to modernize operator-dependent diagnostic workflows while preserving compatibility with existing laboratory practice.

## Acknowledgements

T.E. was partially funded by the Structured Doctoral programme (*Strukturiertes Doktorandenprogramm zum Erwerb des Dr. med. und Dr. rer. nat.*) of Heidelberg University. The authors acknowledge support by the state of Baden-Württemberg through bwHPC and the German Research Foundation (DFG) through grant INST 35/1597-1 FUGG. The authors gratefully acknowledge the data storage service SDS@hd supported by the Ministry of Science, Research and the Arts Baden-Württemberg (MWK) and the German Research Foundation (DFG) through grant INST 35/1803-1 FUGG and INST 35/1804-1 LAGG. The authors acknowledge support by the state of Baden-Württemberg through bwVisu. The study was further supported by funding from the Health + Life Science Alliance Heidelberg Mannheim of the Medical Faculty of the University of Heidelberg. The authors thank Xenia Bengs for critically reviewing the manuscripts Method section. Figures 5A and 6A/B were created using licensed BioRender.

## Data availability statement

A subset of simulated image data (DOI: 10.5281/zenodo.20855843) as well as acquired microscopy data that were used for data analysis in this work (shown in Figure 4) or are referenced by the figures (DOI: 10.5281/zenodo.20963451) and flow cytometry raw data (DOI: 10.5281/zenodo.21396263) are deposited at Zenodo. Source data that the figures were generated from are publicly available at Zenodo as well (DOI: 10.5281/zenodo.20963490).

AlloViewer is available at https://alloviewer.org/.

## Code availability statement

The full source code is available at github (https://github.com/TarikExner/alloviewer). The code to use the API and the code to develop the pipeline and the entirety of this work are part of the same package.

## Ethics Statement

This study was approved by the responsible institutional ethics committee (approval number 2026-3057).

