## Supplementary and Extended Data for "Simulation-Trained Deep Learning for Automated Cell-Based HLA Antibody Assay Interpretation in Pre-Transplant Diagnostics"

### Supplementary Information

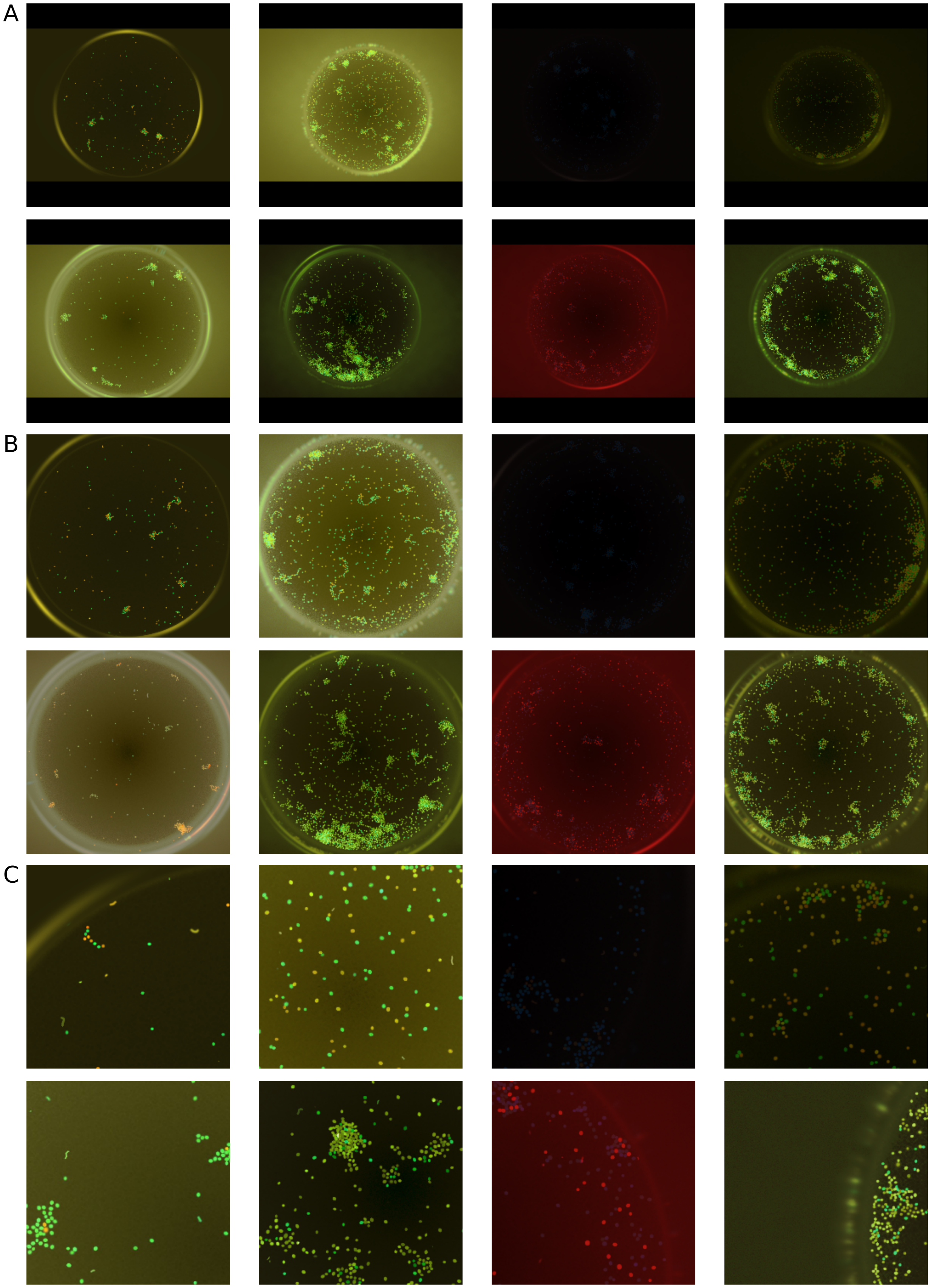
A
B
C

**Supplementary Figure S1. Comparison of image preprocessing strategies for well-level microscopy data.**

**A** Padding and downsizing. Original well images were resized to a fixed shape by adding black padding around the edges, preserving the entire field of view while maintaining the circular well geometry.

**B** Crop-well and resize. The circular well region was cropped from each frame and then resized to a standard resolution, removing surrounding background while retaining the well interior.

**C** Tiling. High-resolution well images were divided into smaller image tiles, each covering a local area of the well, allowing higher effective resolution and detailed visualization of local cell density and morphology. All images are representative examples from the simulated dataset and illustrate how each preprocessing approach affects apparent scale, background content, and visible detail.

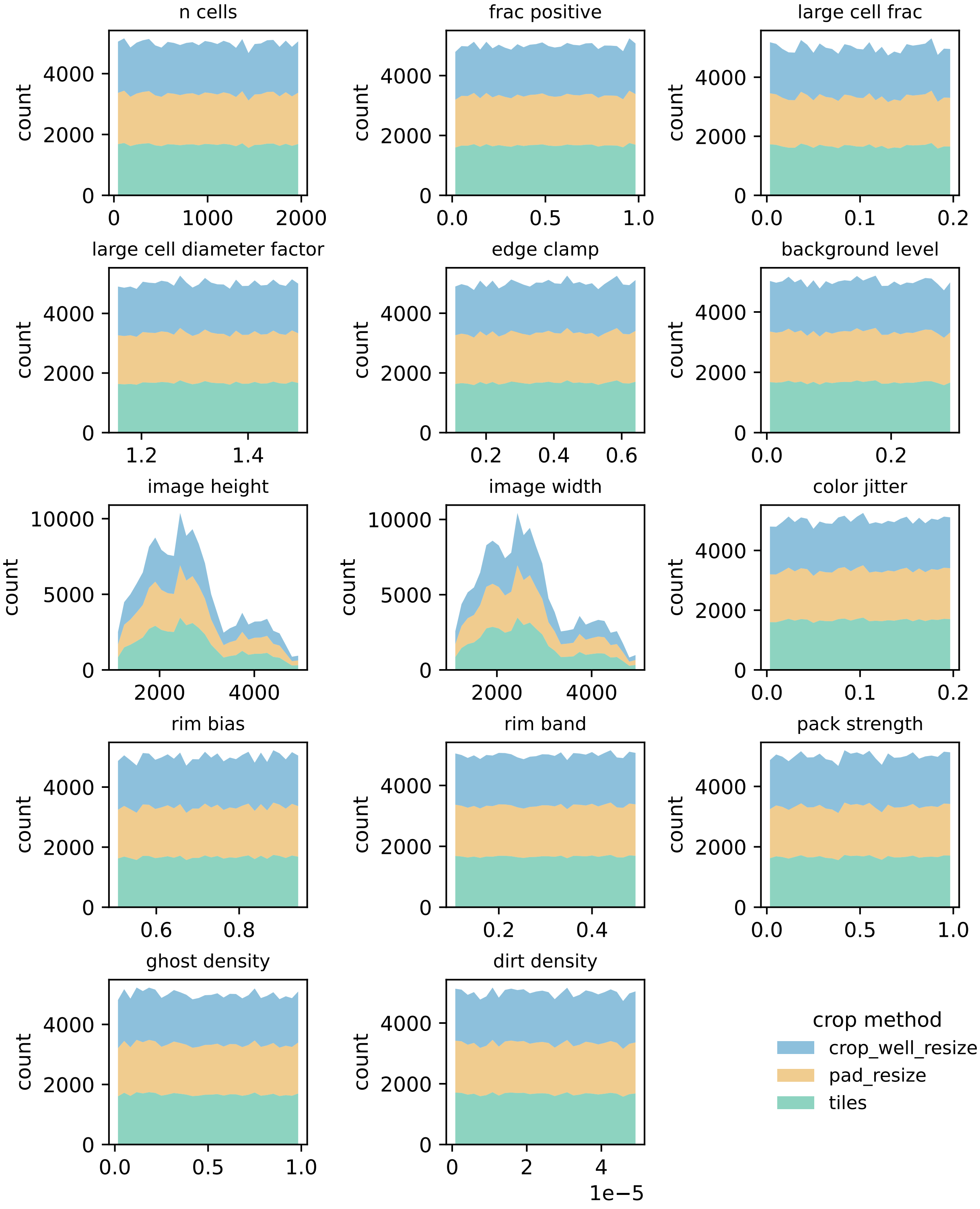
n cells
frac positive
large cell frac
large cell diameter factor
edge clamp
background level
image height
image width
color jitter
rim bias
rim band
pack strength
ghost density
dirt density
count
1e−5
crop method
crop_well_resize
pad_resize
tiles

**Supplementary Figure S2. Coverage of simulation parameters across datasets.** Stacked histograms show the distribution of sampled simulation parameters for the three image crop methods: crop_well_resize, pad_resize and tiles. Colors indicate crop method, and each panel shows the same parameter range across the three groups. The plotted parameters include the number of simulated cells (n_cells), the positive-cell fraction (frac_positive), the fraction of large cells (large_cell_frac), and the size multiplier for large cells (large_cell_diameter_factor). Image geometry is represented by image_height and image_width. The parameter background_level controls the base brightness of the well, while color_jitter controls per-cell color variation. Spatial cell placement is described by rim_bias, rim_band and edge_clamp, which control how strongly cells are biased toward the well rim and how closely they are constrained near the edge. Pack_strength controls the strength of overlap resolution between cells. Artifact-related parameters include ghost_density, which controls the abundance of faint out-of-focus cells outside the main well region, and dirt_density, which controls the amount of small debris inside the well. The similar distributions across crop methods indicate that the simulation parameter space is sampled comparably across datasets.

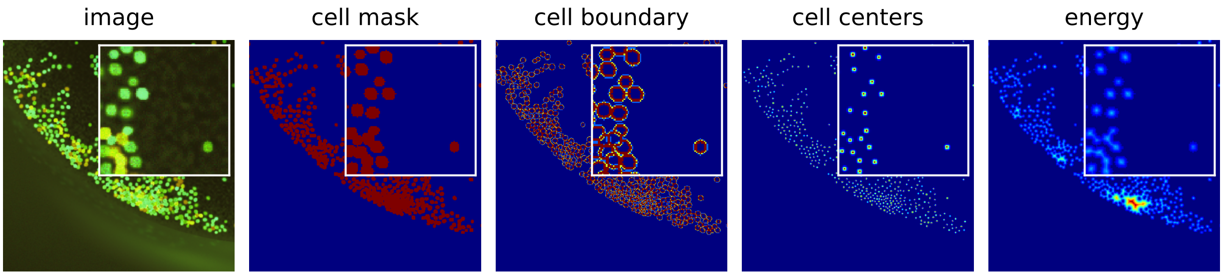
image
cell mask
cell boundary
cell centers
energy

**Supplementary Figure S3. Synthetic training sample and supervision targets.** The first display item in the panel shows a simulated well image. The next four items show the targets used for UNet training. The cell mask is a binary foreground map derived from the instance labels. The cell boundary map highlights a border ring around each cell; it is produced from the instance map and rendered as a narrow band. The cell centers map marks the centroid of each instance and is converted to a small Gaussian heatmap for training. The energy map is the normalized Euclidean distance to the nearest boundary, defined only inside cells and set to zero elsewhere. Insets in all panels zoom into a representative region to show fine detail of the targets.

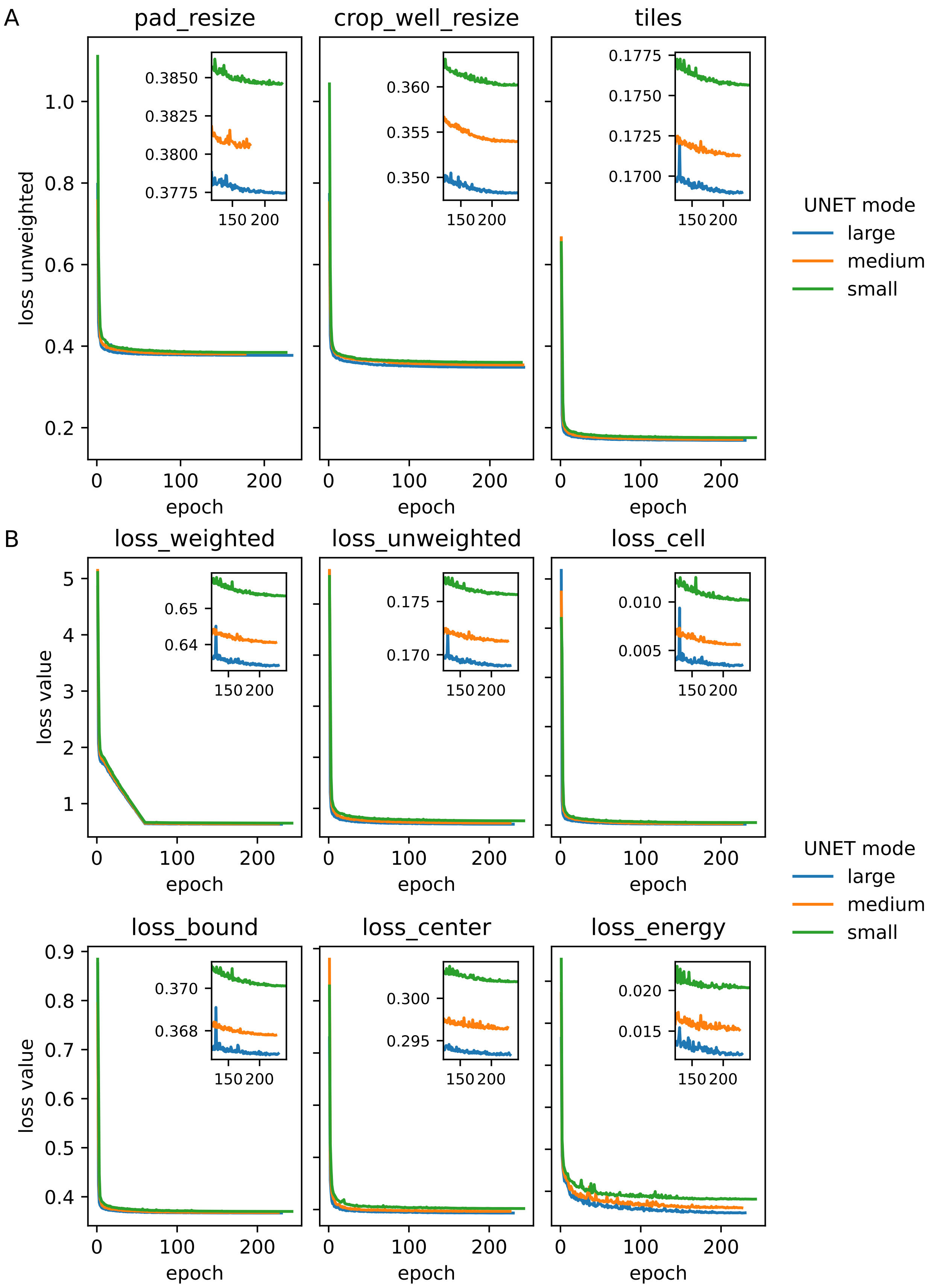
A
pad_resize
crop_well_resize
tiles
loss unweighted
epoch
UNET mode
large
medium
small
B
loss_weighted
loss_unweighted
loss_cell
loss_bound
loss_center
loss_energy
loss value

**Supplementary Figure S4. Validation losses during training for different UNet configurations.**

**A** Validation unweighted loss over training epochs for three data preprocessing modes (pad_resize, crop_well_resize, and tiles). Each curve corresponds to a different UNet size (small, medium, or large). Insets show a magnified view of the final training phase. All three panels share the same y-axis for direct comparison.

**B** Validation losses for the tiles configuration, separated by individual loss components. Only the leftmost panels display the y-axis labeled “loss value.” The loss_weighted curve represents the complete training objective, combining all task-specific losses with additional regularization terms (boundary-exclusion and anti-halo penalties). The loss_unweighted curve is the unweighted mean of the individual head losses and serves as the model selection criterion. The loss_cell term measures segmentation accuracy of the main cell mask using a mixture of binary cross-entropy and Dice loss. The loss_bound term quantifies errors in predicting the soft boundary probability map. The loss_center term evaluates the accuracy of detecting cell centroids through a combined BCE and mean squared error formulation, emphasizing positive samples with a positional weighting. The loss_energy term reflects the regression error of an energy map within the segmented cells, calculated as a combination of L1 and MSE losses. Lower values indicate better performance across all panels. Insets in each plot highlight fine-scale improvements toward convergence.

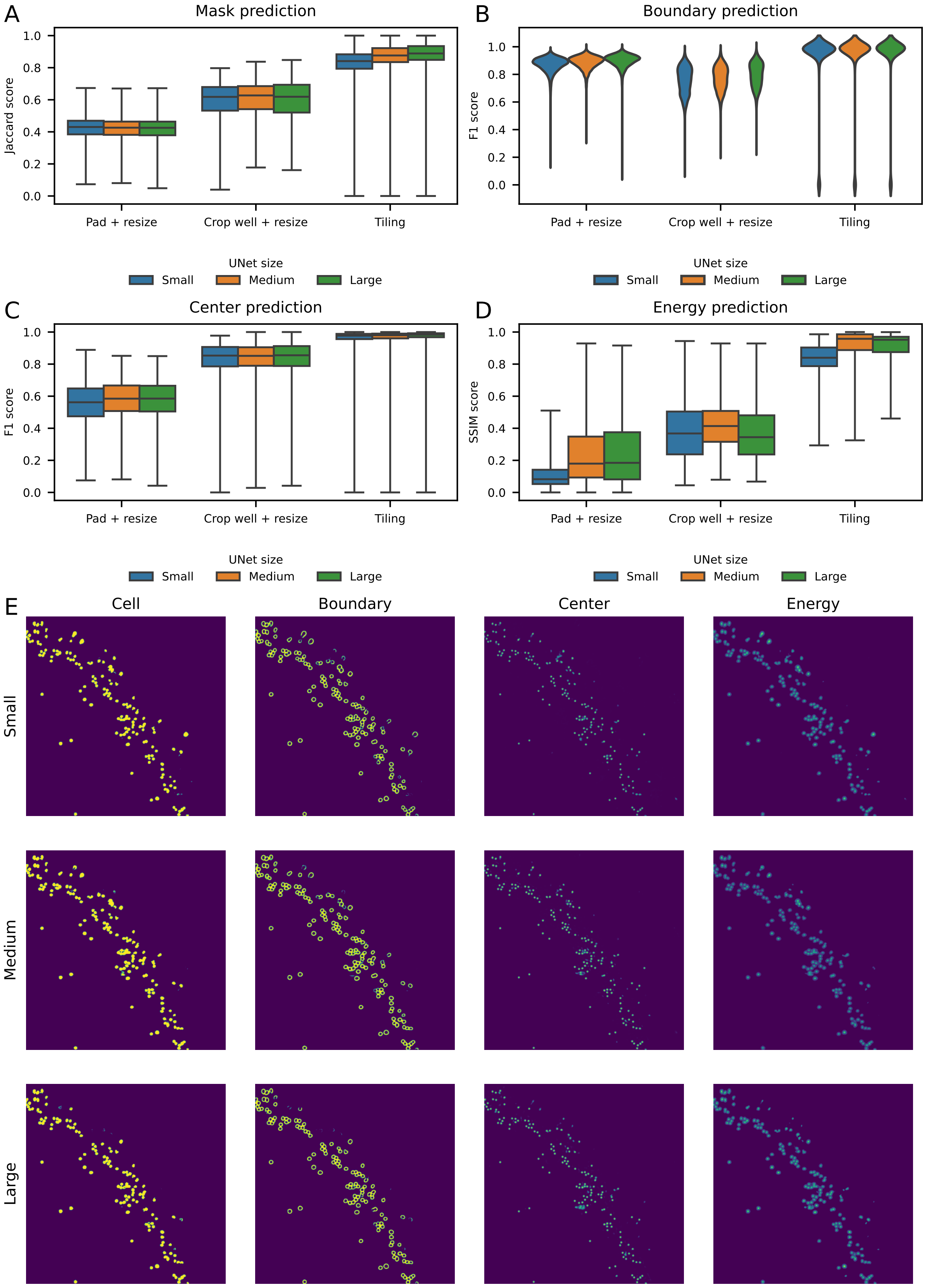
A
Mask prediction
Jaccard score
Pad + resize
Crop well + resize
Tiling
B
Boundary prediction
F1 score
UNet size
Small
Medium
Large
C
Center prediction
D
Energy prediction
SSIM score
E
Cell
Boundary
Center
Energy
Small
Medium
Large

**Supplementary Figure S5. Comparison of UNet sizes and dataset preprocessing strategies.**

Performance of three UNet architectures (large, medium, small) was evaluated across three data preprocessing strategies for obtaining the final dataset: pad_resize, crop_well_resize, and tiling.

**A** Mask prediction: Segmentation quality measured by the Jaccard (IoU) score.

**B** Boundary prediction: F1 score of predicted boundary pixels versus ground truth.

**C** Center prediction: Accuracy of predicted cell centers measured by the F1 score.

**D** Energy prediction: Structural similarity (SSIM) between predicted and ground truth energy maps.

**E** Example tiles from real microscopy images for each UNet size and image prediction (raw probabilities: cell, bound, center, energy); the outputs of the three UNet sizes are visually largely indiscernible, showing that UNet size has only a minor effect.

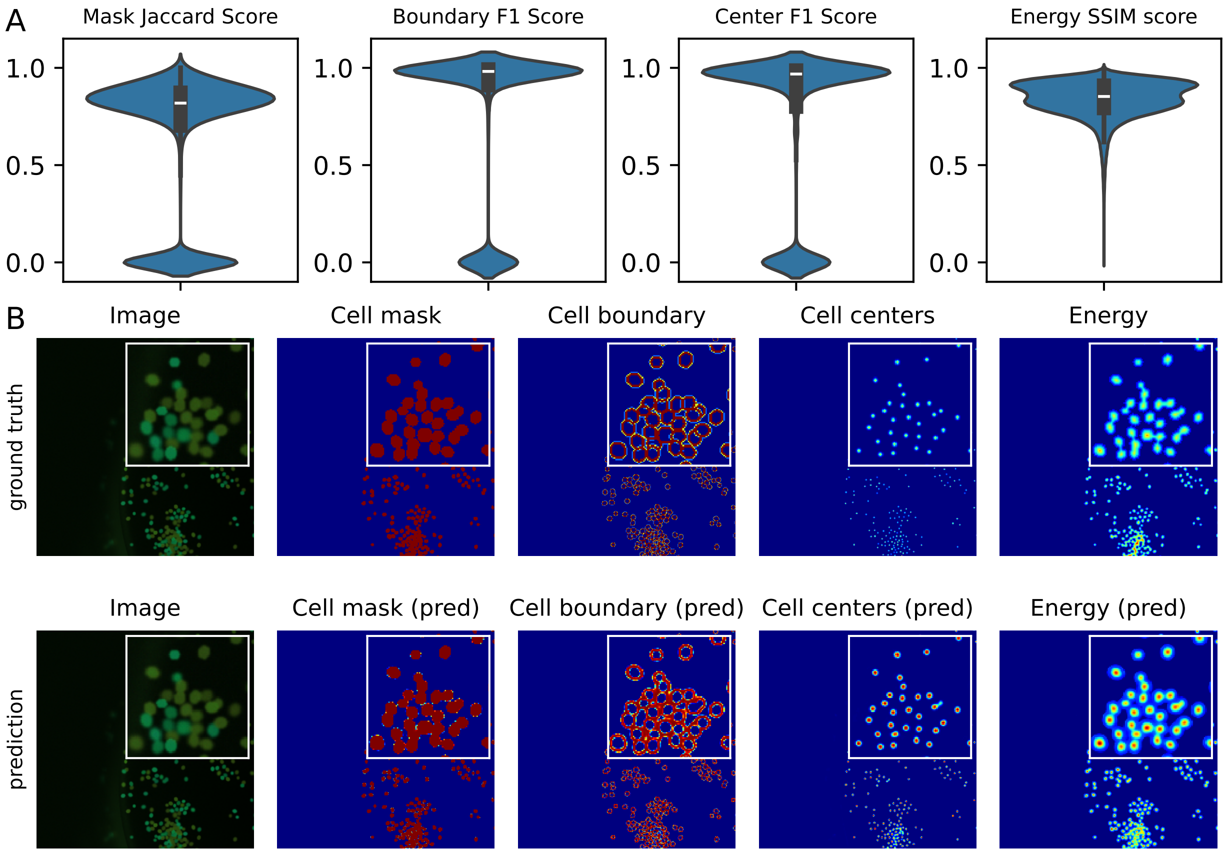
A
Mask Jaccard Score
Boundary F1 Score
Center F1 Score
Energy SSIM score
1.0
0.5
0.0
B
Image
Cell mask
Cell boundary
Cell centers
Energy
ground truth
Image
Cell mask (pred)
Cell boundary (pred)
Cell centers (pred)
Energy (pred)
prediction

**Supplementary Figure S6. UNet performance on simulated cell images.**

**A** Violin plots of segmentation quality on the simulated test set. The *Mask Jaccard score* (IoU) measures how much the predicted cell mask overlaps with the true mask. The *Boundary F1 score* measures how well the predicted edges follow the true cell edges within a small tolerance. The *Center F1 score* measures how well cell centers are detected using one-to-one matching between predicted and true centers. The *Energy SSIM score* measures how similar the predicted and true per-cell energy maps are inside cells; values close to 1 mean very strong to perfect agreement. Across all four metrics, the UNet reaches high scores on the simulated data. (n=5000).

**B** Example simulated field of view. Top row: input image with the four ground truth maps (cell mask, boundary, centers, and energy). Bottom row: the corresponding UNet predictions. Insets zoom into a small region and show that predicted masks, centers, and energy closely follow the ground truth. pred, predicted.

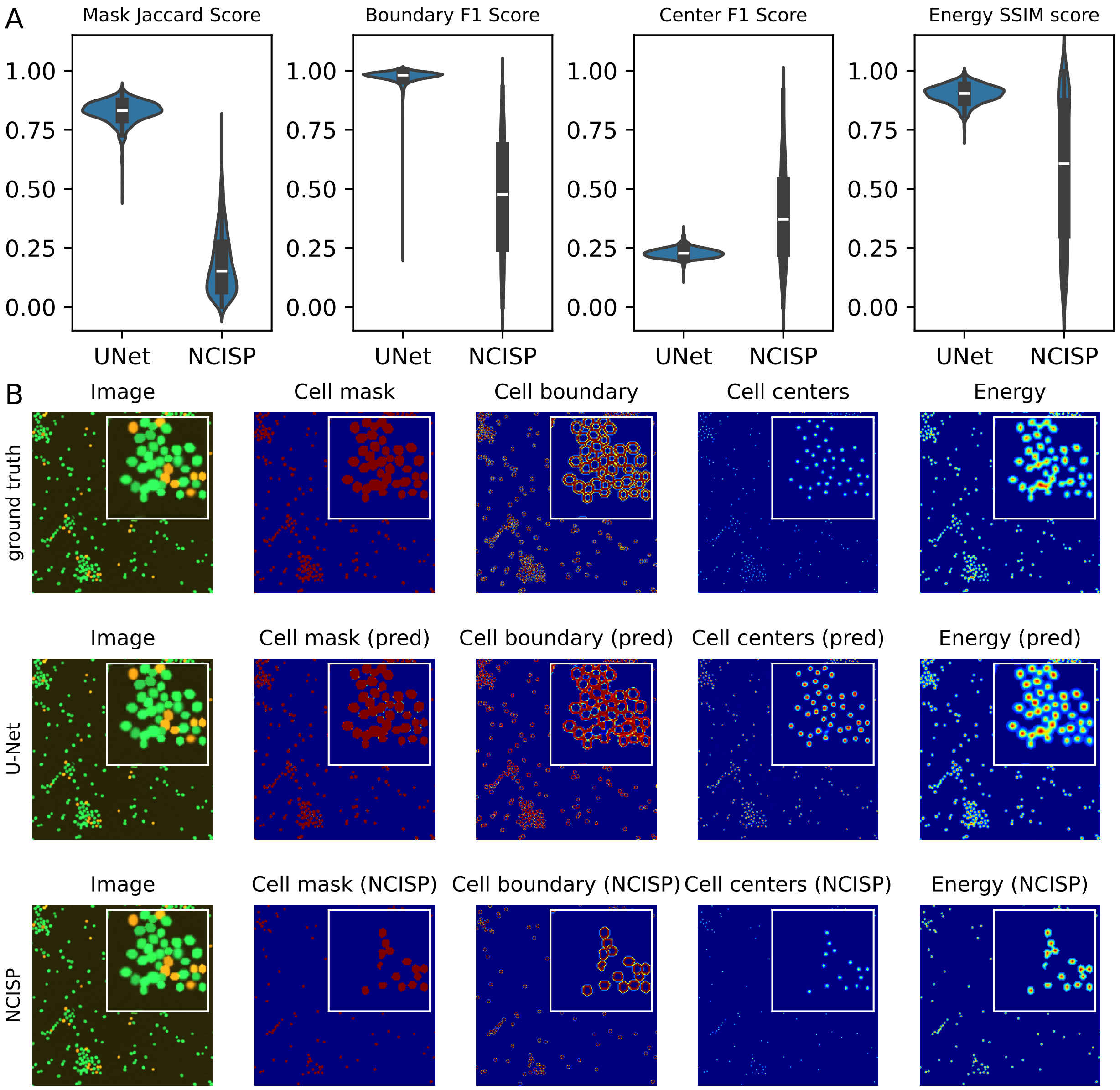
A
Mask Jaccard Score
Boundary F1 Score
Center F1 Score
Energy SSIM score
1.00
0.75
0.50
0.25
0.00
UNet
NCISP
B
Image
Cell mask
Cell boundary
Cell centers
Energy
ground truth
Cell mask (pred)
Cell boundary (pred)
Cell centers (pred)
Energy (pred)
U-Net
Cell mask (NCISP)
Cell boundary (NCISP)
Cell centers (NCISP)
Energy (NCISP)
NCISP

**Supplementary Figure S7. Comparison of UNet and NCISP segmentations on simulated cell images.**

**A** Violin plots comparing segmentation quality between the UNet model and NCISP across four metrics on the simulated test dataset. The Mask Jaccard score (IoU) measures mask overlap. The Boundary F1 score evaluates how well the detected cell edges follow the true boundaries. The Center F1 score reflects how accurately individual cell centers are detected. The Energy SSIM score measures similarity between predicted and true per-cell energy maps. Across all metrics, the UNet achieves consistently higher values than NCISP.

**B** Visual comparison for a representative simulated field of view. *Top row*: Ground truth image and masks. *Middle row*: UNet predictions. *Bottom row*: NCISP segmentations. Insets zoom into a representative crowded region. Although scaled to the same bit-depth, simulated images are only poorly segmented using the NCISP pipeline, suggesting that the parameters are not matched well for the simulated intensity distribution that reflects inter- and intra-laboratory acquisition differences.

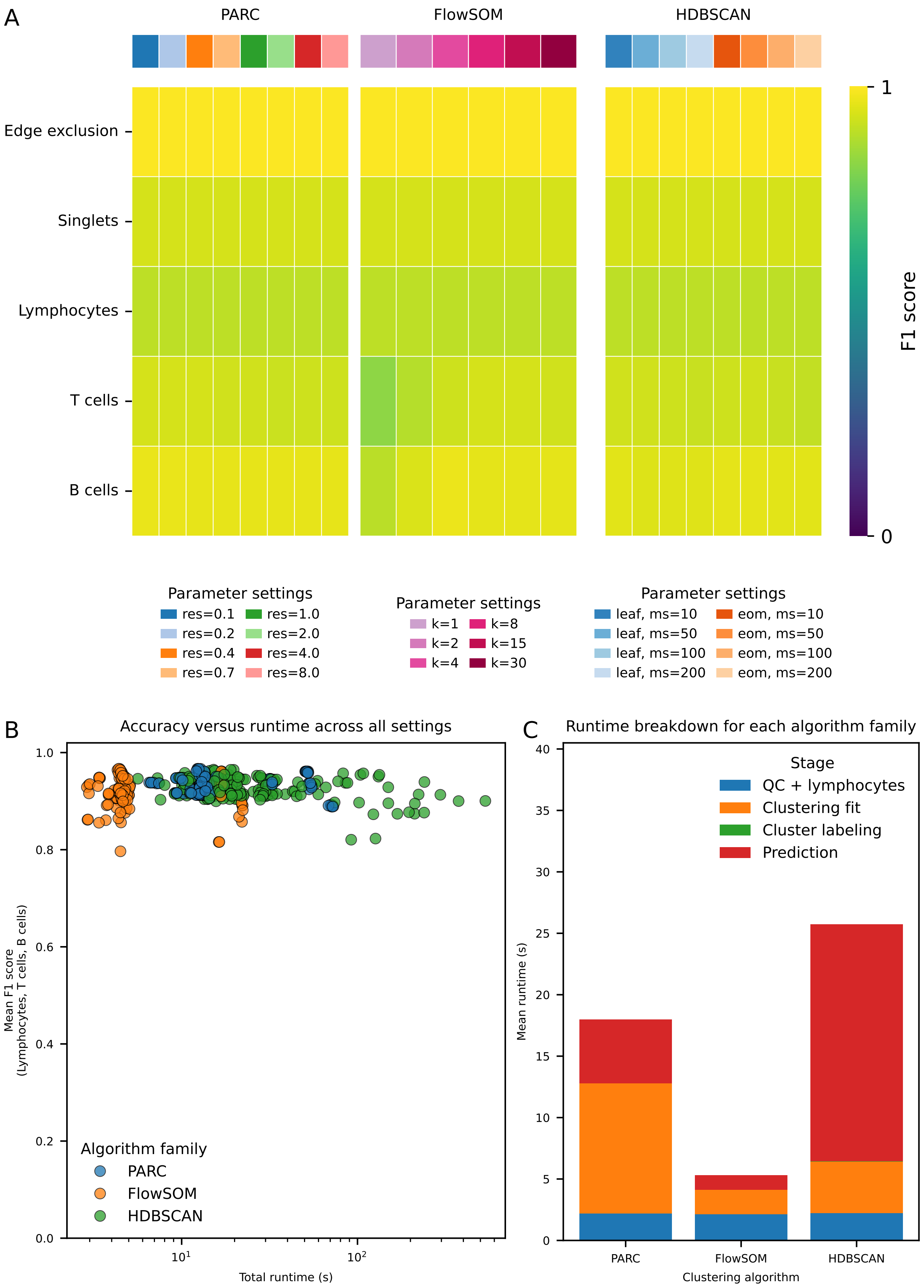
A
PARC
FlowSOM
HDBSCAN
Edge exclusion
Singlets
Lymphocytes
T cells
B cells
F1 score
1
0
Parameter settings
res=0.1
res=0.2
res=0.4
res=0.7
res=1.0
res=2.0
res=4.0
res=8.0
Parameter settings
k=1
k=2
k=4
k=8
k=15
k=30
Parameter settings
leaf, ms=10
leaf, ms=50
leaf, ms=100
leaf, ms=200
eom, ms=10
eom, ms=50
eom, ms=100
eom, ms=200
B
Accuracy versus runtime across all settings
Mean F1 score (Lymphocytes, T cells, B cells)
Total runtime (s)
Algorithm family
PARC
FlowSOM
HDBSCAN
C
Runtime breakdown for each algorithm family
Mean runtime (s)
Clustering algorithm
Stage
QC + lymphocytes
Clustering fit
Cluster labeling
Prediction
PARC
FlowSOM
HDBSCAN

**Supplementary Figure S8. Clustering parameter choices have only modest effects on gating accuracy, whereas runtime differs substantially across algorithm families and settings.**

**A** Mean F1-scores across all FCXM experiments (n=23) for each tested parameter setting of PARC, FlowSOM, and HDBSCAN, shown separately for each gating target. Rows indicate gating targets and columns indicate parameter settings, with parameter identities marked by the colored annotation bars above each heatmap and listed in the legends below. Color encodes agreement with manual ground truth gating. res, PARC resolution parameter; k, FlowSOM number of metaclusters; leaf/eom, cluster selection methods with their respective settings.

**B** Accuracy-versus-runtime comparison across all tested parameter settings. Each point represents one experiment and one parameter setting, plotted by total runtime and mean F1-score across the biologically relevant gates Lymphocytes, T-cells, and B-cells. Points are colored by the algorithm family.

**C** Mean runtime breakdown for the best-performing parameter setting within each algorithm family. Bars are stacked by pipeline stage (QC; Quality Control) plus Lymphocyte gating, clustering fit, cluster labeling and prediction, and averaged across experiments. FlowSOM achieved similar accuracy to PARC and HDBSCAN at substantially lower runtime, whereas PARC and HDBSCAN required longer computation, with prediction accounting for a large share of total runtime.

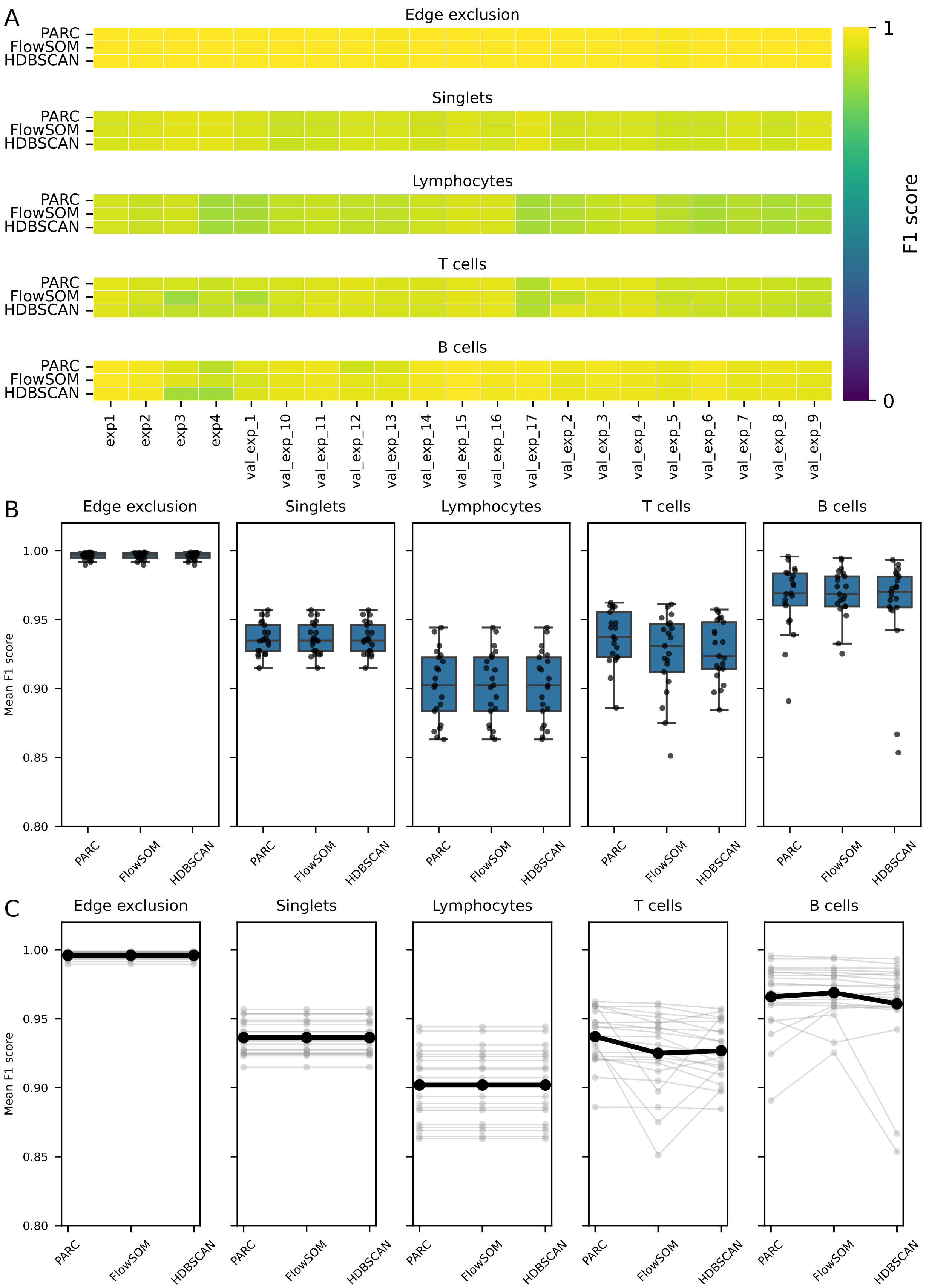
A
Edge exclusion
Singlets
Lymphocytes
T cells
B cells
PARC
FlowSOM
HDBSCAN
exp1
exp2
exp3
exp4
val_exp_1
val_exp_10
val_exp_11
val_exp_12
val_exp_13
val_exp_14
val_exp_15
val_exp_16
val_exp_17
val_exp_2
val_exp_3
val_exp_4
val_exp_5
val_exp_6
val_exp_7
val_exp_8
val_exp_9
F1 score
1
0
B
Mean F1 score
1.00
0.95
0.90
0.85
0.80
C
Mean F1 score

**Supplementary Figure S9. Best-performing clustering settings show highly similar accuracy across experiments, with only small differences between algorithm families.** **A** Experiment-level mean F1-scores for the best-performing parameter setting within each algorithm family (PARC, FlowSOM, HDBSCAN), shown separately for each gating target. Rows indicate algorithm families, columns indicate experiments, and color encodes agreement with manual ground truth gating. Best settings were selected based on mean performance across the biologically relevant gates Lymphocytes, T-cells, and B-cells.
**B** Distribution of experiment-level mean F1-scores for these best-performing settings. Boxes indicate the interquartile range, center lines mark the median, whiskers show the range excluding outliers, and points represent individual experiments (n=23).
**C** Paired experiment-level comparison of the same best-performing settings across algorithm families. Thin gray lines connect the same experiment across PARC, FlowSOM, and HDBSCAN, and the thick black line shows the mean across experiments. Edge exclusion and singlet gating were highly consistent across methods, whereas larger between-experiment spread was observed for lymphocyte, T-cell, and B-cell identification. Overall, the three algorithm families showed closely matched performance across experiments (n=23). exp, experiment; val_exp, validation experiment.

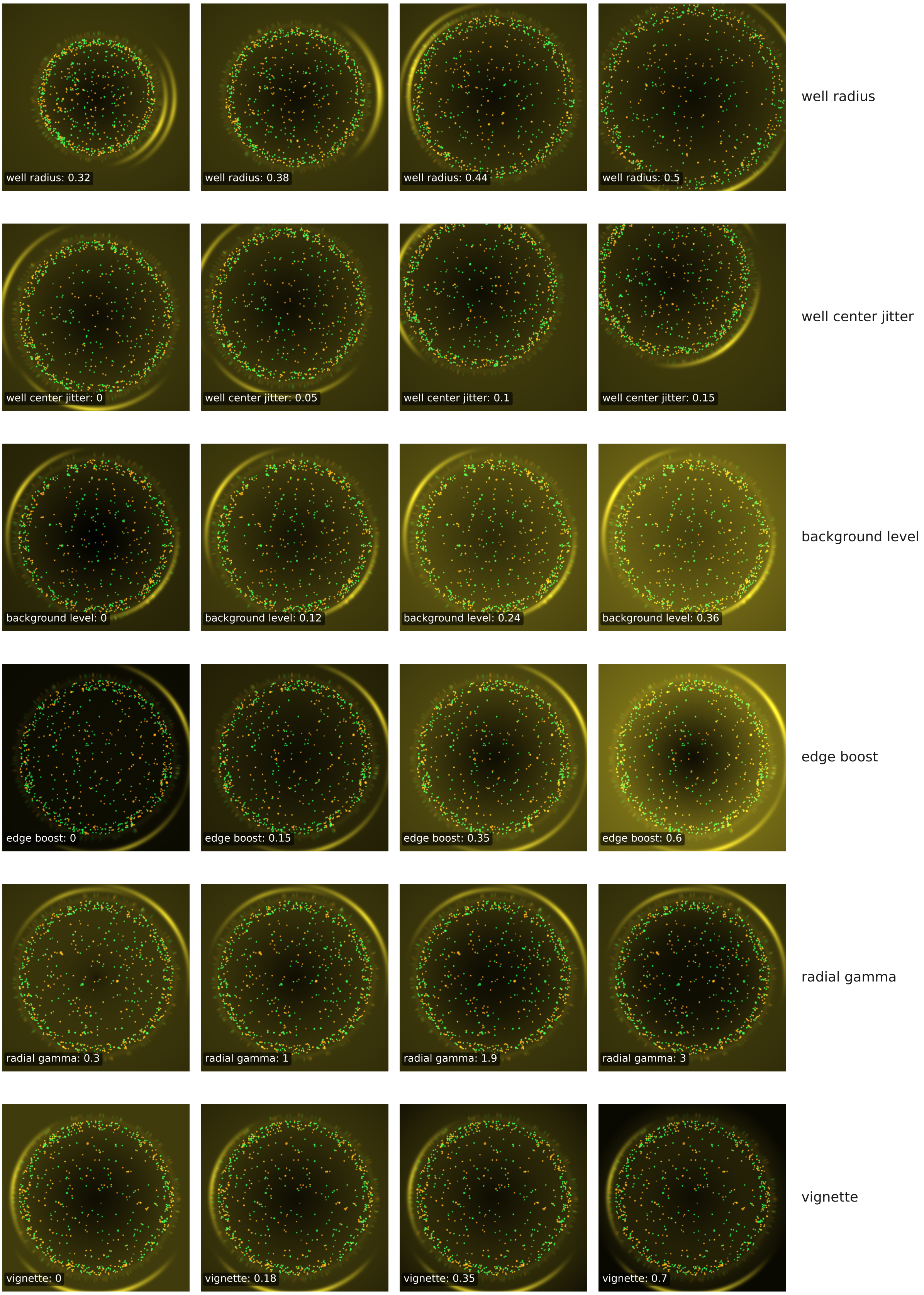
well radius: 0.32
well radius: 0.38
well radius: 0.44
well radius: 0.5
well radius
well center jitter: 0
well center jitter: 0.05
well center jitter: 0.1
well center jitter: 0.15
well center jitter
background level: 0
background level: 0.12
background level: 0.24
background level: 0.36
background level
edge boost: 0
edge boost: 0.15
edge boost: 0.35
edge boost: 0.6
edge boost
radial gamma: 0.3
radial gamma: 1
radial gamma: 1.9
radial gamma: 3
radial gamma
vignette: 0
vignette: 0.18
vignette: 0.35
vignette: 0.7
vignette

**Extended Data Figure 1. Parameter effects on well geometry and background intensity.** Synthetic fluorescence images showing the effect of varying well radius, well-center jitter, background level, radial edge boost, radial intensity exponent, and vignette strength. In each row, only the indicated parameter was varied while the remaining simulator settings were held constant.

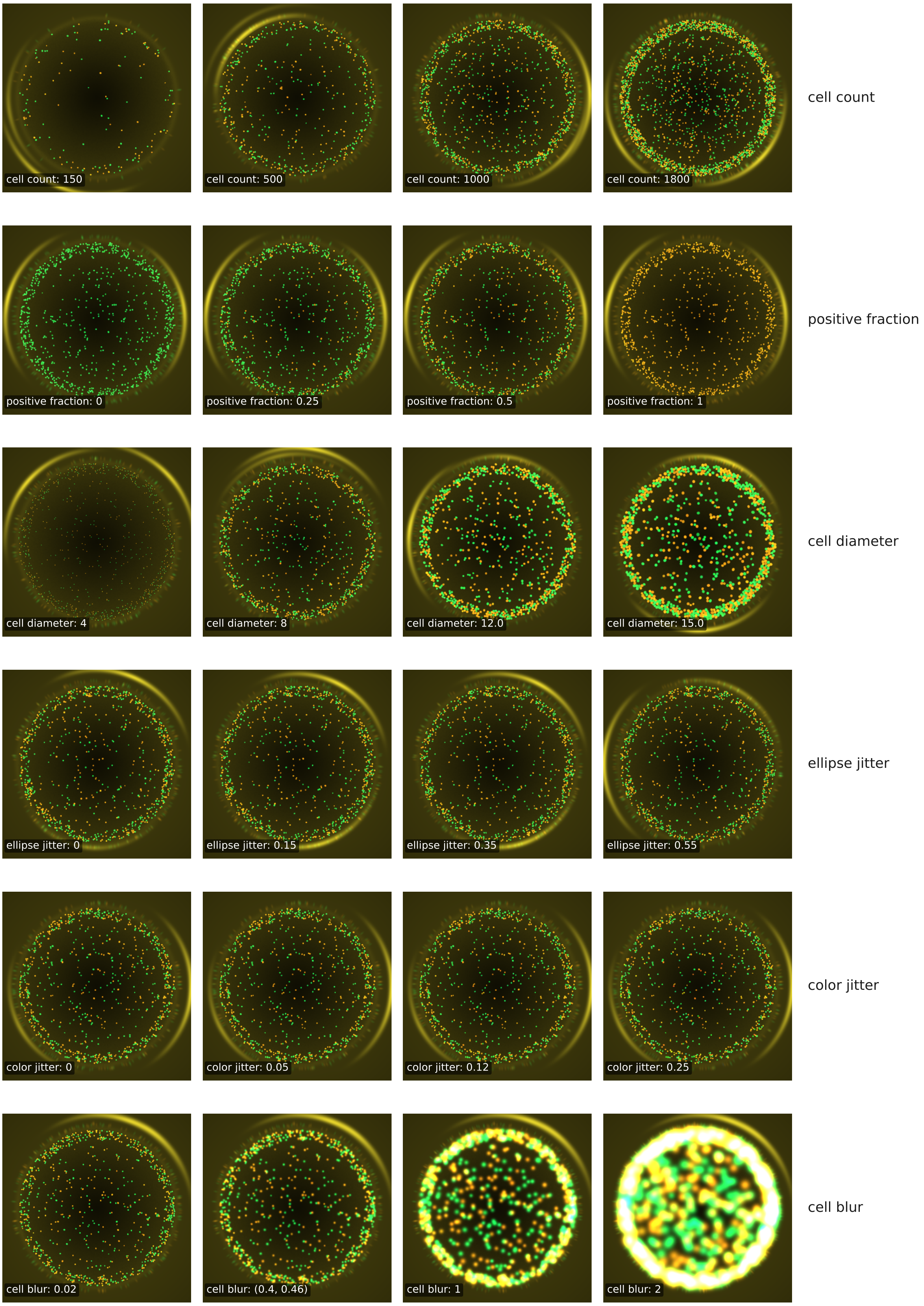
cell count: 150
cell count: 500
cell count: 1000
cell count: 1800
cell count
positive fraction: 0
positive fraction: 0.25
positive fraction: 0.5
positive fraction: 1
positive fraction
cell diameter: 4
cell diameter: 8
cell diameter: 12.0
cell diameter: 15.0
cell diameter
ellipse jitter: 0
ellipse jitter: 0.15
ellipse jitter: 0.35
ellipse jitter: 0.55
ellipse jitter
color jitter: 0
color jitter: 0.05
color jitter: 0.12
color jitter: 0.25
color jitter
cell blur: 0.02
cell blur: (0.4, 0.46)
cell blur: 1
cell blur: 2
cell blur

**Extended Data Figure 2. Parameter effects on cell appearance.**
Synthetic images showing changes in cell number, positive-cell fraction, cell diameter, ellipticity, color jitter, and blur. These sweeps illustrate how the simulator controls cell density, class balance, apparent cell size and shape, channel variation, and focus-related smoothing.

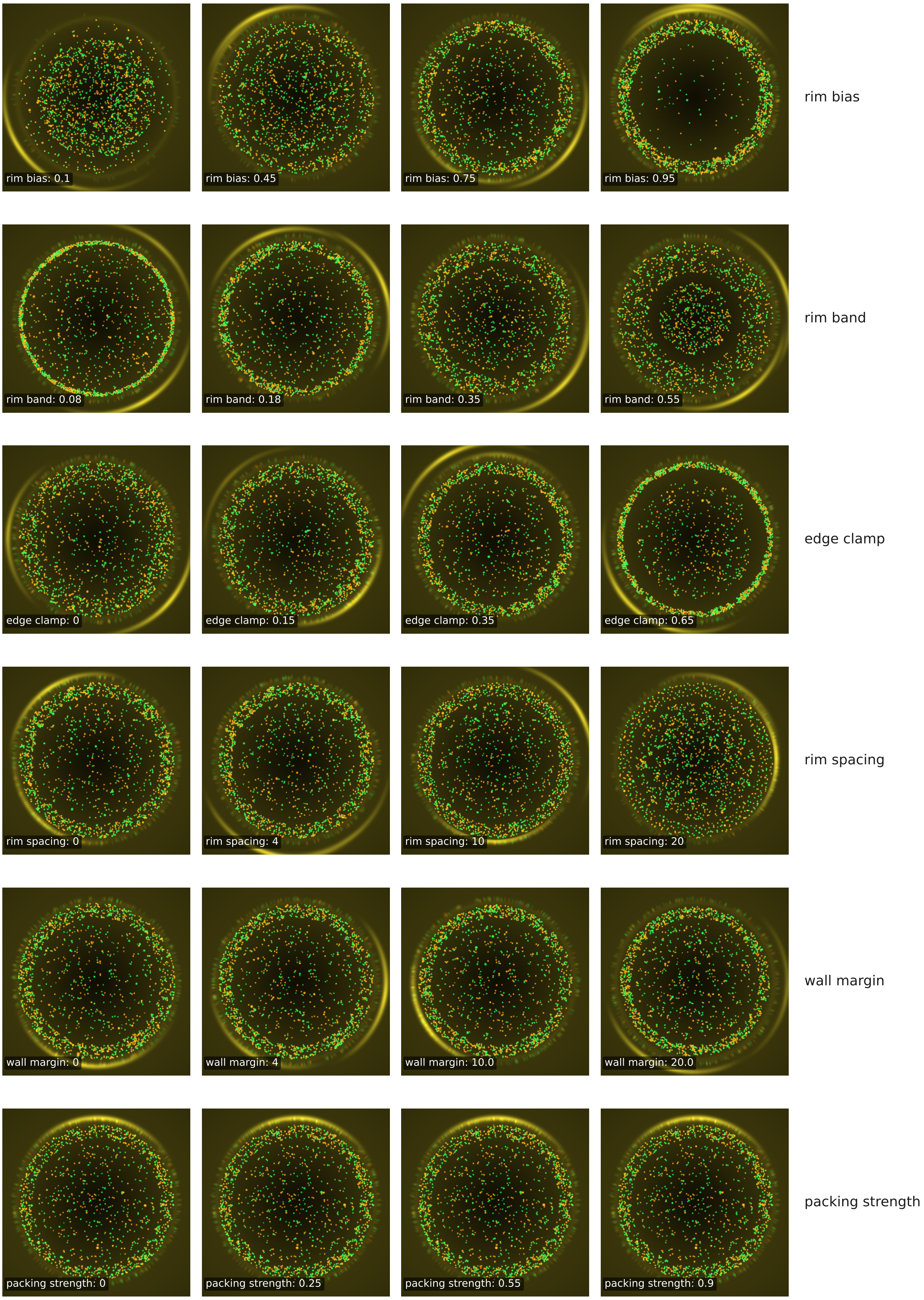
rim bias: 0.1
rim bias: 0.45
rim bias: 0.75
rim bias: 0.95
rim bias
rim band: 0.08
rim band: 0.18
rim band: 0.35
rim band: 0.55
rim band
edge clamp: 0
edge clamp: 0.15
edge clamp: 0.35
edge clamp: 0.65
edge clamp
rim spacing: 0
rim spacing: 4
rim spacing: 10
rim spacing: 20
rim spacing
wall margin: 0
wall margin: 4
wall margin: 10.0
wall margin: 20.0
wall margin
packing strength: 0
packing strength: 0.25
packing strength: 0.55
packing strength: 0.9
packing strength

**Extended Data Figure 3. Parameter effects on radial cell placement and packing.** Synthetic images showing how rim bias, rim-band width, edge clamping, rim spacing, wall margin, and packing strength affect the spatial distribution of cells inside the well. The examples show transitions from more uniform placement to stronger accumulation near the well rim and altered local cell spacing.

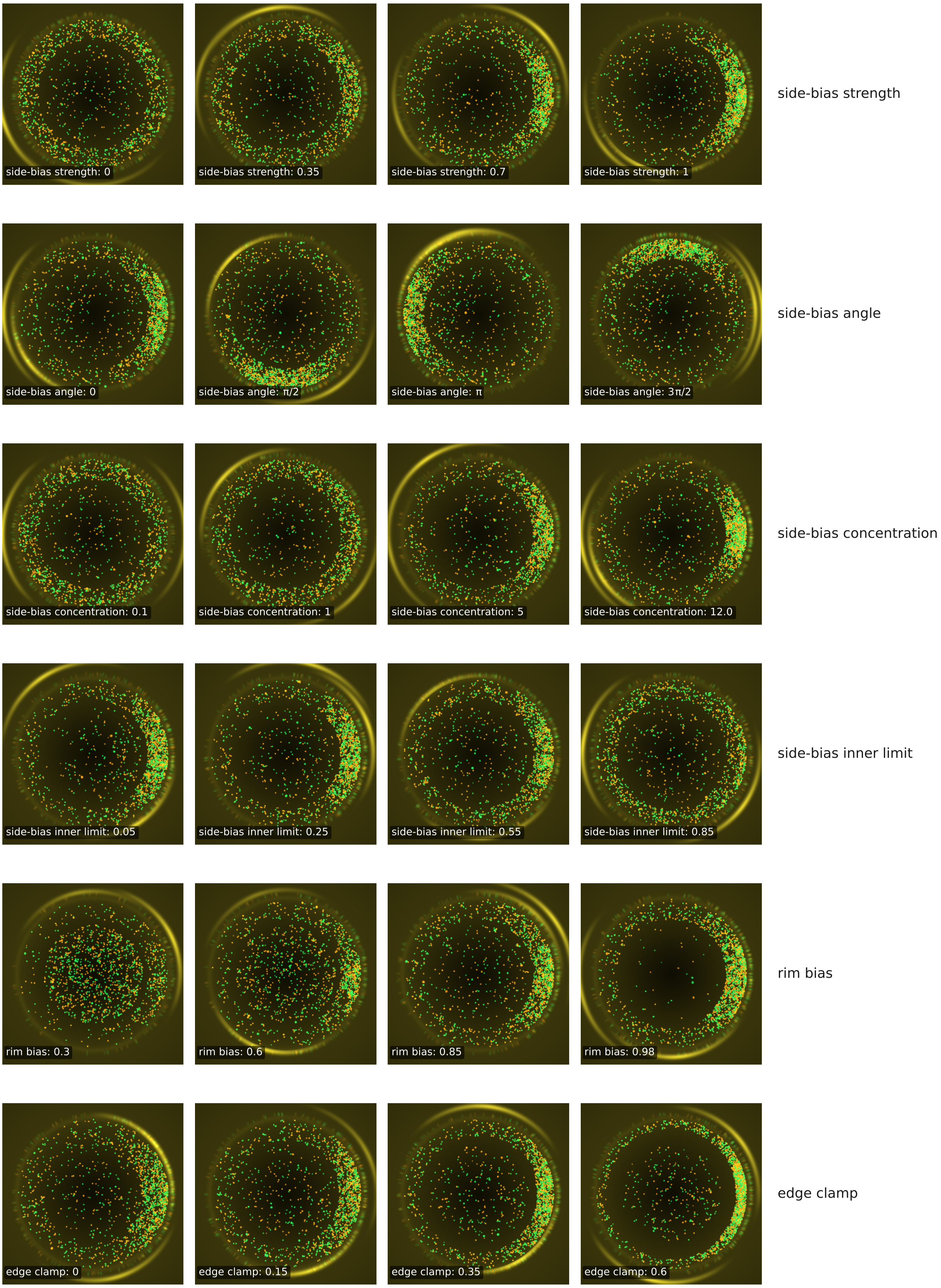
side-bias strength: 0
side-bias strength: 0.35
side-bias strength: 0.7
side-bias strength: 1
side-bias strength
side-bias angle: 0
side-bias angle: π/2
side-bias angle: π
side-bias angle: 3π/2
side-bias angle
side-bias concentration: 0.1
side-bias concentration: 1
side-bias concentration: 5
side-bias concentration: 12.0
side-bias concentration
side-bias inner limit: 0.05
side-bias inner limit: 0.25
side-bias inner limit: 0.55
side-bias inner limit: 0.85
side-bias inner limit
rim bias: 0.3
rim bias: 0.6
rim bias: 0.85
rim bias: 0.98
rim bias
edge clamp: 0
edge clamp: 0.15
edge clamp: 0.35
edge clamp: 0.6
edge clamp

**Extended Data Figure 4. Parameter effects on one-sided placement bias.** Synthetic images showing asymmetric cell accumulation produced by the side-bias parameters. The sweeps vary side-bias strength, angular direction, angular concentration, inner radial limit, rim bias, and edge clamping, demonstrating how the simulator can reproduce directional pile-up patternss near the well edge.

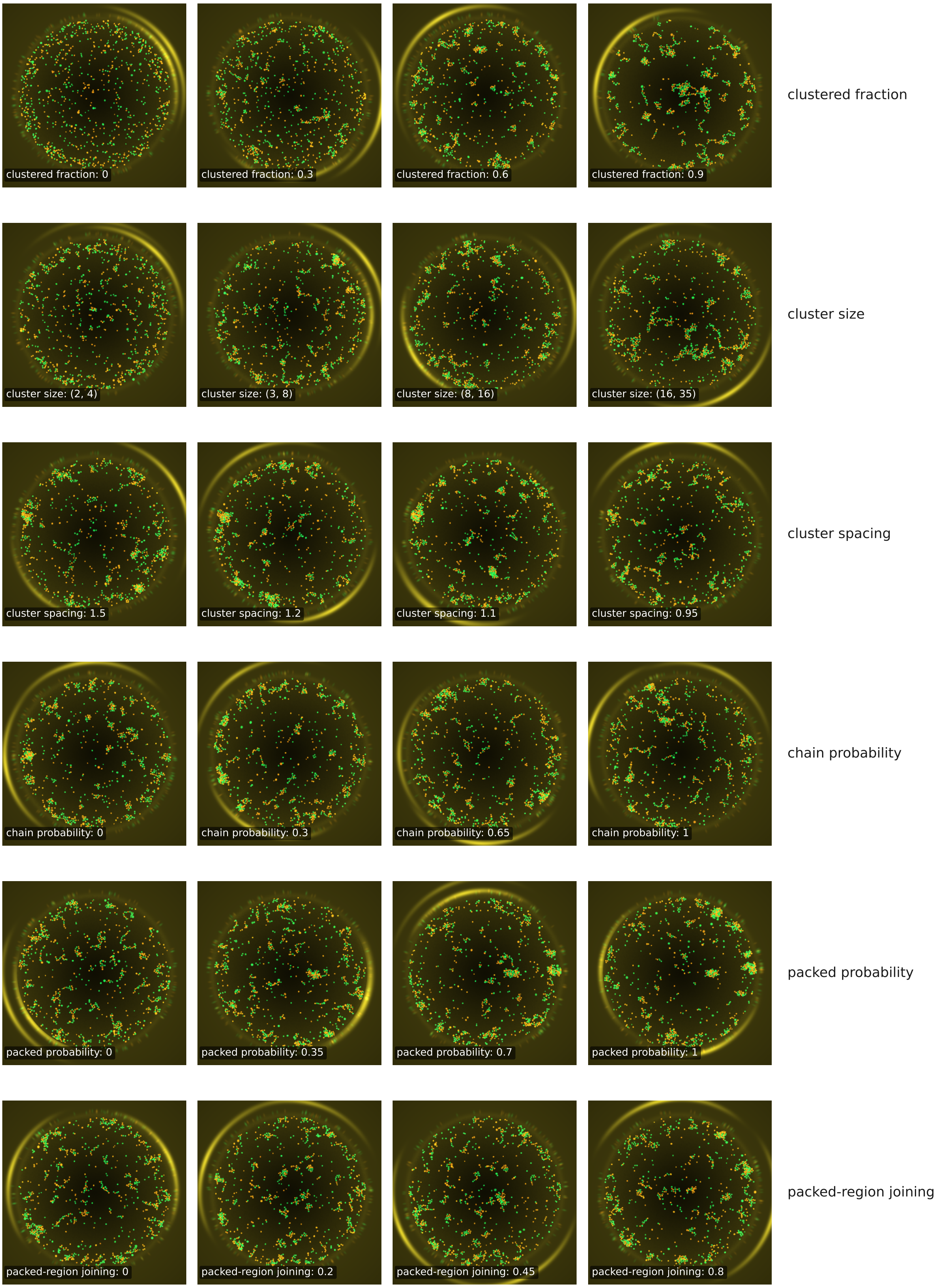
clustered fraction: 0
clustered fraction: 0.3
clustered fraction: 0.6
clustered fraction: 0.9
clustered fraction
cluster size: (2, 4)
cluster size: (3, 8)
cluster size: (8, 16)
cluster size: (16, 35)
cluster size
cluster spacing: 1.5
cluster spacing: 1.2
cluster spacing: 1.1
cluster spacing: 0.95
cluster spacing
chain probability: 0
chain probability: 0.3
chain probability: 0.65
chain probability: 1
chain probability
packed probability: 0
packed probability: 0.35
packed probability: 0.7
packed probability: 1
packed probability
packed-region joining: 0
packed-region joining: 0.2
packed-region joining: 0.45
packed-region joining: 0.8
packed-region joining

**Extended Data Figure 5. Parameter effects on clustered cell placement.** Synthetic images showing explicit clustered placement of cells. The rows vary the fraction of clustered cells, cluster-size range, cluster spacing, probability of chain-like clusters, probability of packed clusters, and probability that packed clusters join larger packed regions.

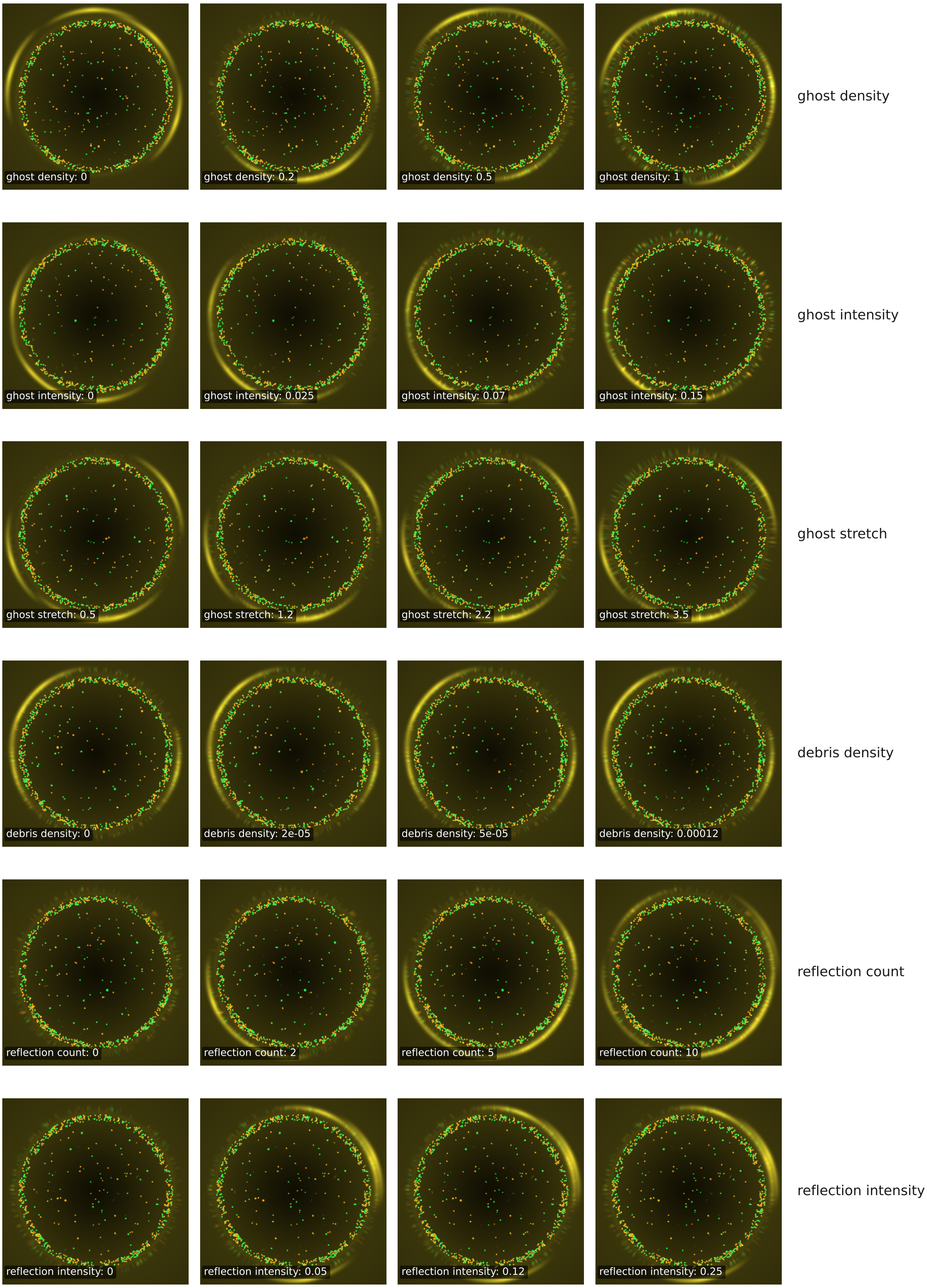
ghost density: 0
ghost density: 0.2
ghost density: 0.5
ghost density: 1
ghost density
ghost intensity: 0
ghost intensity: 0.025
ghost intensity: 0.07
ghost intensity: 0.15
ghost intensity
ghost stretch: 0.5
ghost stretch: 1.2
ghost stretch: 2.2
ghost stretch: 3.5
ghost stretch
debris density: 0
debris density: 2e-05
debris density: 5e-05
debris density: 0.00012
debris density
reflection count: 0
reflection count: 2
reflection count: 5
reflection count: 10
reflection count
reflection intensity: 0
reflection intensity: 0.05
reflection intensity: 0.12
reflection intensity: 0.25
reflection intensity

**Extended Data Figure 6. Parameter effects on image-only artifacts.**
Synthetic images showing artifacts that are rendered into the image but excluded from ground-truth cell labels. The rows vary ghost-cell density (cells outside the main focus-plane of the well), ghost-cell intensity, ghost elongation, debris density, reflection count, and reflection intensity.
